# Developmental Glymphatic Dysfunction Underlies Excitation/Inhibition Imbalance and Psychosis Vulnerability in 22q11.2 Deletion Syndrome

**DOI:** 10.1101/2025.08.29.25334716

**Authors:** Alessandro Pascucci, Silas Forrer, Corrado Sandini, Valentina Mancini, Yasser Alemán-Gómez, Stephan Eliez, Farnaz Delavari

## Abstract

**Background:** Impairment of the glymphatic system may contribute to atypical brain development and increased vulnerability to psychiatric conditions such as psychosis. In particular, disrupted glymphatic efficiency may affect neurochemical homeostasis during critical maturational windows, leading to structural and circuit-level alterations. However, its role in early neurodevelopmental trajectories remains largely unexplored.

**Methods:** We combined longitudinal diffusion tensor imaging (DTI) and magnetic resonance spectroscopy (MRS) in individuals with 22q11.2 deletion syndrome (22q11DS), a condition associated with elevated psychosis risk. Glymphatic function was estimated using the DTI-ALPS index, based on both manual and automated ROI placement. Excitation/inhibition ratio was assessed in the right hippocampus via CSF-corrected Glx and GABA levels.

**Results:** ALPS index was significantly reduced in 22q11DS compared to controls (p = 0.017), especially in the right hemisphere. Individuals with positive psychotic symptoms (PPS+) showed a divergent developmental trajectory, failing to exhibit the age-related ALPS increase seen in PPS− (group x age interaction: p = 0.009). In a subset with spectroscopy data (n = 39), lower ALPS predicted higher Glx/GABA ratio in the right hippocampus (p = 0.002).

**Conclusions:** These findings provide *in vivo* evidence that glymphatic dysfunction emerges early and follows atypical developmental trajectories in those at risk for psychosis. Impaired clearance is also associated with excitatory/inhibitory imbalance. This dysfunction may represent a novel pathway contributing to psychosis vulnerability and a potential target for early intervention.

## 1. INTRODUCTION

Understanding how deviations in normal brain development result in psychiatric illness requires an integrative framework encompassing genetic vulnerabilities, environmental exposures, and the biological systems that mediate their interaction. Among these systems, the glymphatic clearance network has emerged as a promising but underexplored point of convergence, potentially linking diverse risk factors to disrupted brain homeostasis during critical periods of development. This perivascular network, known as the glymphatic system (GS), drives cerebrospinal fluid (CSF) into brain parenchyma through astrocytic aquaporin-4 (AQP4) channels and directs interstitial by-products, including potassium, excess glutamate, cytokines, and misfolded proteins, toward venous and meningeal lymphatic pathways (1–4). Efficient glymphatic flow is essential for synaptic remodeling, metabolite recycling, and overall circuit maturation (5). Its disruption can amplify excitotoxicity, oxidative stress, and neuroinflammatory processes that have been repeatedly implicated in psychosis pathophysiology (6–8). Thus, glymphatic dysfunction may represent a mechanistic pathway from disrupted neurophysiology to pathobiology of psychosis. This is supported by postmortem, CSF, and PET imaging studies showing elevated inflammatory mediators and evidence of microglia activation in individuals with schizophrenia and in those at clinical high risk who later developed psychosis (9–14). In parallel, magnetic resonance spectroscopy (MRS) studies document imbalances in major excitatory and inhibitory neurotransmitters in the cortex and hippocampus (15–20). These imbalances correlate with progressive hippocampal volume loss, possibly via neurotoxic mechanisms, contributing to emergence of psychosis (21–23). While such abnormalities may arise from intrinsic dysregulation of neurotransmitter synthesis and reuptake, impaired clearance represents an underexplored contributor that may allow harmful metabolites to accumulate and disrupt synaptic homeostasis.

Human imaging studies have already linked inefficient glymphatic flow to neurodegenerative diseases, cognitive decline, traumatic brain injury, and mood disorders (24–29). These findings suggest that impaired clearance affects normal brain function, allowing neurotoxic by-products to accumulate and ultimately leading to structural and functional brain alterations. Glymphatic dysfunction has also been reported in neurodevelopmental conditions such as autism spectrum disorder and schizophrenia, although most of these investigations have been cross-sectional, limiting insight into how such alterations evolve over time (30–32) . The glymphatic network itself matures postnatally, with increasing evidence indicating progressive organization of perivascular pathways throughout adolescence and into early adulthood (33–35). However, the developmental trajectories of glymphatic function, and their interaction with mechanisms known to contribute to psychosis, remain largely uncharacterized. Addressing this gap is critical, as enhancing glymphatic clearance during key developmental windows, for example with ameliorating sleep quality or supporting blood brain barrier integrity, may represent a viable therapeutic opportunity before irreversible circuit damage occurs(36–38).

22q11.2 deletion syndrome (22q11DS) offers a valuable model for studying glymphatic system development. This microdeletion is associated with up to a 40-fold increased risk for schizophrenia and other neurodevelopmental disorders, making it a relevant clinical and biological model for psychiatric risk (39–42). Several structural components of the glymphatic system appear to be directly affected in 22q11DS. Cellular and animal studies have reported compromised blood–brain barrier (BBB) integrity (43–45), altered astrocyte maturation (46,47), and impaired ependymal cilia function (48), all crucial elements for efficient glymphatic clearance. Furthermore, patients with 22q11DS are often identified at birth due to somatic features of the syndrome, long before psychiatric symptoms emerge, thereby enabling longitudinal investigations of early-stage neurobiological alterations (49).

To capture glymphatic system function *in vivo*, we leveraged diffusion tensor imaging analysis along the perivascular space, known as the ALPS index. This imaging approach provides a reproducible and non-invasive proxy for glymphatic clearance efficiency, previously validated in studies of aging and neurodegenerative disorders (50–53).

In the largest longitudinal cohort of individuals with 22q11.2DS studied to date, we mapped age-related changes in glymphatic system efficiency using the ALPS index and tested whether impaired clearance was linked to the emergence of positive psychotic symptoms. Additionally, we investigated a subgroup of deletion carriers with available MRS datasets to examine the relationship between glymphatic clearance efficiency and excitation/inhibition imbalance, a neurochemical alteration already implicated both in 22q11DS population and in psychosis more broadly (15,21). Through this integrative approach, we aimed to determine if and when glymphatic dysfunction first emerges during development, how it intersects with established pathogenic pathways, and where intervention efforts might best be targeted to support healthy brain maturation.

## 2. MATERIAL AND METHODS

### 2.1. Participants

This study included longitudinal assessments of 85 individuals with a confirmed diagnosis of 22q11.2 Deletion Syndrome (22q11DS) (143 scans; mean age: 18.92 ± 6.36 years; 43 males and 42 females) and 83 HCs (115 scans; mean age: 16.28 ± 6.42 years; 40 males and 43 females), all recruited as part of the Swiss 22q11DS longitudinal cohort. Participants were aged at scan between 5 and 35 years and contributed a total of 258 diffusion tensor imaging (DTI) scans across multiple time points. Details of MRI data acquisition and processing are provided in the **Supplementary Methods**.

HCs were recruited from siblings of deletion carriers and through an open call from the Geneva state school system. They were screened to exclude any current or past neurological or psychiatric diagnoses, learning disabilities, history of prematurity, or use of psychotropic medications. In the 22q11DS group, the microdeletion was confirmed using quantitative fluorescent polymerase chain reaction (qfPCR). Written informed consent was obtained from all participants and/or their legal guardians. The study protocol was approved by the Institutional Review Board of Geneva University School of Medicine and conducted in accordance with the Declaration of Helsinki.

### 2.2. Clinical assessment

All participants with 22q11DS underwent clinical evaluation by an expert psychiatrist (SE), including a semi-structured interview and the Structured Interview for Psychosis-Risk Syndromes (SIPS). In line with previous publications of the same group, presence of clinically significant positive psychotic symptoms was defined as a score ≥ 3 on any of the five positive symptom items (P1–P5) at any time-points. This criterion was chosen as it has been verified to be one of the criteria for ultra-high risk status in this population (54–58).

### 2.3. Data Acquisition

All participants underwent MRI on a 3T Siemens Trio scanner using standardized acquisition protocols. Diffusion-weighted images were acquired with 30 directions (b = 1000 s/mm²), and MEGA-PRESS ¹H-MRS was used to quantify GABA+ and Glx in the right hippocampus. Voxel placement was guided by individual T1-weighted scans. For full acquisition parameters and preprocessing details, please refer to the **Supplementary Methods**.

### 2.4. ALPS Index Method

The ALPS index was computed following previously published methodology to quantify direction-specific diffusivity along perivascular spaces, serving as an indirect marker of glymphatic clearance function (50,51,59,60). Diffusion tensor components along the three orthogonal axes (Dxx, diffusivity in the right–left direction; Dyy, diffusivity in the anterior–posterior direction; Dzz, diffusivity in the inferior–superior direction) were extracted in native space using a custom pipeline described in **Supplementary Methods**. Spherical regions of interest (ROIs, 4 mm diameter) were manually placed bilaterally at the level of the lateral ventricles, targeting projection and association white matter fibers, whose principal directions are predominantly along the z- and y-axes, respectively. ROIs were positioned adjacent to medullary veins, which course along the x-axis, presumed to align with PVSs, while avoiding areas of fiber crossing in the same direction (52). The ALPS index was computed separately for each hemisphere using the following formula:

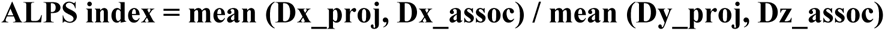

Where Dx_proj denotes diffusivity in the x-direction (right–left) within projection fiber ROIs, Dx_assoc denotes diffusivity in the x-direction within association fiber ROIs, Dy_proj denotes diffusivity in the y-direction (anterior–posterior) within projection fiber ROIs, and Dz_assoc denotes diffusivity in the z-direction (inferior–superior) within association fiber ROIs. Higher values reflect greater diffusivity along PVs relative to perpendicular directions, consistent with more efficient glymphatic clearance. A bilateral ALPS index was obtained by averaging the left and right hemisphere values.

### 2.5. Automated ROI positioning and replication

To ensure reproducible and anatomically consistent ROI placement across participants, an automated pipeline was developed to project standard-space ROIs into individual diffusion space. Four bilateral ROIs, targeting the superior corona radiata (SCR) and superior longitudinal fasciculus (SLF), were defined in MNI space using the JHU-ICBM-FA-2mm atlas. Non-linear registration was then used to map these ROIs to each subject’s native space for subject-specific ALPS index computation. A visual overview of the ROI placement in MNI and subject space is shown in **Figures 1b and 1c**, respectively. Full technical details of the registration and transformation procedures are provided in the **Supplementary Methods**.

**Figure 1.**
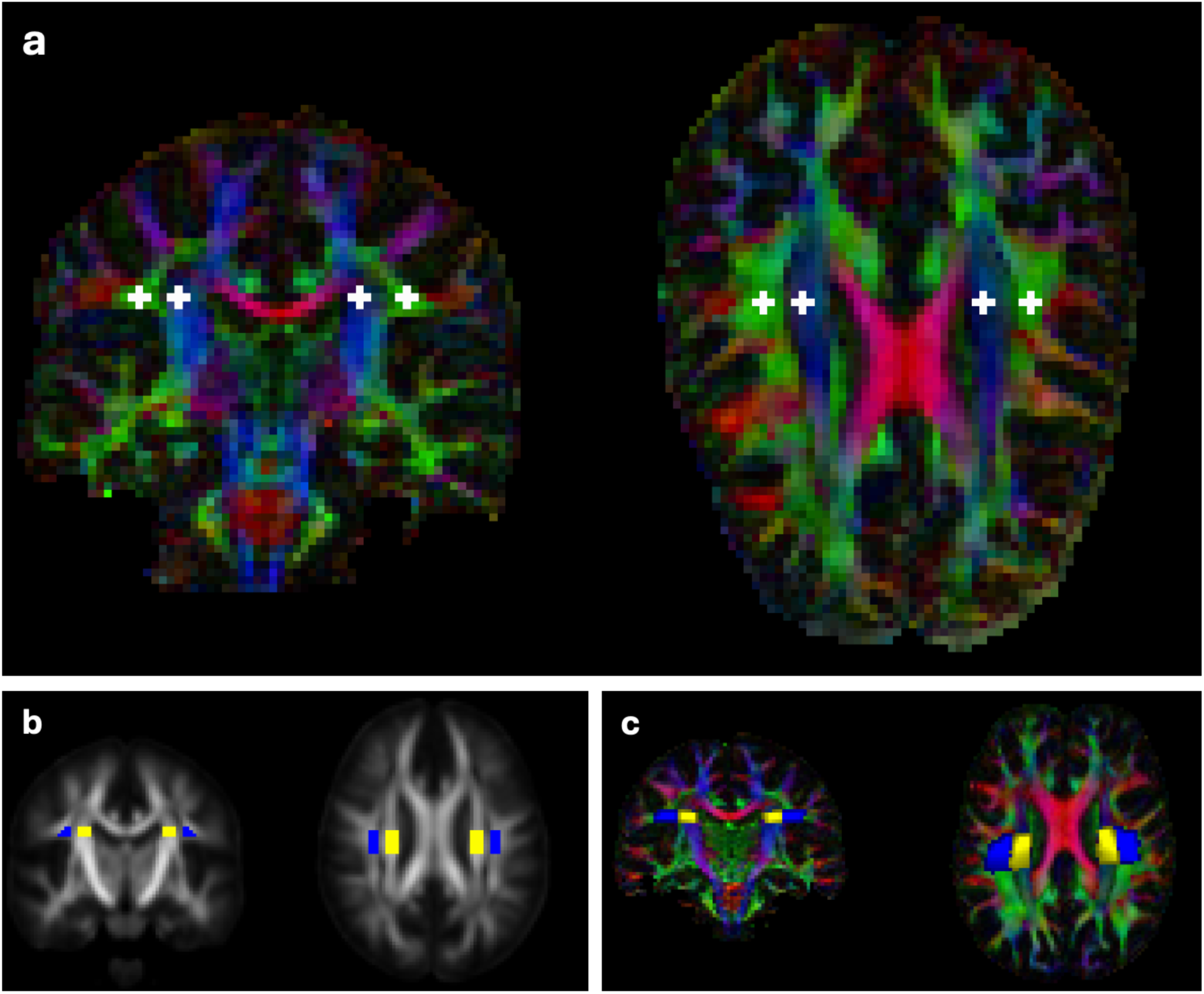
ALPS Index Calculation (a) Manual ROI Placement Approach. Schematic representation of the manual method for calculating the ALPS index. ROIs were manually placed in projection and association fiber areas adjacent to the lateral ventricles, guided by colored FA maps. **(b) Automated Method: ROI Placement on MNI template:** JHU-ICBM-FA-2mm atlas with 4 ROI templates: Yellow: Left and Right projection ROIs, Blue: Left and Right association ROIs **(c) Automated Method: ROIs after transforming into subject space (randomly picked subject):** Subject specific colormap obtained from the FA map plus 4 ROIs warped: Yellow: Left and Right projection ROIs, Blue: Left and Right association ROIs.

### 2.6. Statistical Analysis

Left and right ALPS indices were computed separately, and outliers were removed using the interquartile range (IQR ±1.5) method, applied independently by group (HC and 22q11DS). Bilateral mean ALPS values were derived from scans with valid indices in both hemispheres. Psychosis-stratified analyses included 78 individuals with 22q11DS who had valid SIPS assessments. Based on positive symptom scores (P1–P5 ≥ 3), 38 were classified as psychosis-positive (PPS+), and 40 as psychosis-negative (PPS−), contributing 123 scans in total. Outliers were removed within each subgroup as above. Detailed sample characteristics and scan counts after exclusions are reported in the **Supplementary Methods**. Statistical analyses accounted for the nested structure of the longitudinal dataset, which included 258 DTI scans across development in both individuals with 22q11DS and HCs. Linear mixed-effects models were used to evaluate group differences and developmental trajectories, with ALPS index as the dependent variable. Models included fixed effects for diagnostic group, age (mean-centered), sex, and the group-by-age interaction, along with a subject-level random intercept and random slope for age to account for repeated measures. Mixed-effects models were implemented in Python (using the statsmodels package) with fixed effects for group, age (mean-centred), sex, and the group-by-age interaction, and subject-level random effects to account for repeated measures. The same modelling approach was applied to ALPS indices derived from both the manual and automated ROI methods. Statistical significance was set at p < 0.05.

### 2.7. MRS Processing and Analysis

In a subset of 39 individuals with 22q11DS (mean age = 21.6 ± 6.9 years; 25 males), both highquality spectroscopy and diffusion data were available. Proton magnetic resonance spectroscopy (^1^H-MRS) data were processed using Gannet, a widely used toolbox for GABA-edited MRS analysis, to quantify GABA+, Glx, and unsuppressed water signals, and to compute Glx/GABA ratios as a proxy for excitatory/inhibitory (E/I) balance (11,21,61,62).

Tissue segmentation from T1-weighted images enabled correction for partial volume and modeling of tissue-related relaxation properties. Data quality was assessed, and spectra exceeding predefined fit error thresholds were excluded. Further methodological details, including modeling parameters and quality control thresholds, are provided in the **Supplementary Methods**. Linear regressions were performed in Python using the statsmodels package (OLS function) to test the association between ALPS index (predictor) and Glx/GABA ratio (outcome), controlling for age and sex. Analyses were conducted using left, right, and bilateral ALPS values, with outliers excluded via the interquartile range method (IQR ±1.5). A single outlier scan was excluded due to a low right ALPS value; no Glx/GABA outliers were detected. All data were cross-sectional (i.e., one scan per subject).

## 3. RESULTS

### 3.1. Early Glymphatic Dysfunction in 22q11DS

To evaluate group-level differences in trajectory of glymphatic clearance efficiency, we compared ALPS index values between individuals with 22q11DS and HC over age. Mixed-effects models revealed a significant reduction in the bilateral average ALPS index in the 22q11DS group compared to controls (*p* = 0.017; **Figure 2a**). This effect was also significant in the right hemisphere (*p* = 0.022; **Figure 2c**). The left hemisphere showed a slightly reduced ALPS index in 22q11DS, but the difference did not reach statistical significance (*p* = 0.253, **Figure 2b**).

**Figure 2.**
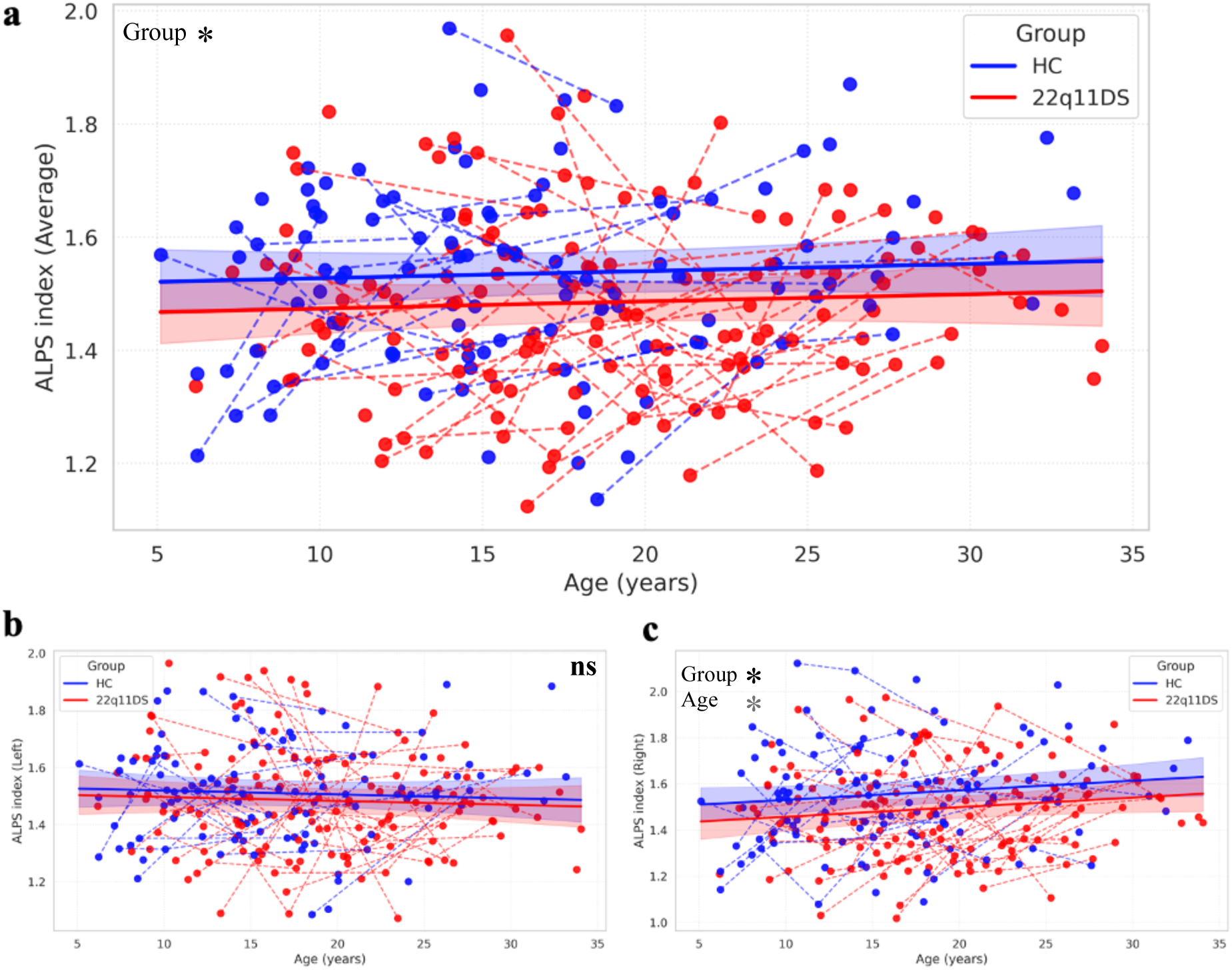
Age-related trajectories of ALPS-index in 22q11.2DS and HC based on manual ROI placement. **(a)** Average ALPS index plotted against age in individuals with 22q11.2 deletion syndrome (22q11DS; red) and healthy controls (HC; blue). Each dot represents a scan; dashed lines connect longitudinal scans from the same subject. Shaded areas represent 95% confidence intervals of the fitted linear mixed-effects models. A significant group difference in the average ALPS index was observed, with lower values in the 22q11DS group (p = 0.017). **(b)** ALPS index in the left hemisphere showed a non-significant trend toward lower values in 22q11DS compared to controls (p = 0.253). **(c)** Right hemisphere ALPS index demonstrated a significant reduction in the 22q11DS group (p = 0.022), alongside a significant effect of age (p = 0.037), indicating a developmental modulation specific to this region. All models included age (mean-centered), group, sex, and age-by-group interaction as fixed effects, and subject-level random slopes for age to account for repeated measures. Black asterisks indicate significant group differences, and grey asterisks indicate significant age effects. *p < 0.05, **p < 0.01, ***p < 0.001, ****p < 0.0001; ns = not significant.

The group × age interaction was not significant in any model, indicating parallel developmental trajectories across groups. Age was not a significant predictor in the left or bilateral ALPS models, suggesting stability of these measures across development. By contrast, age was a significant predictor of right ALPS values (p = 0.037; **Figure 2c**). As the group × age interaction was not significant, this effect was observed in both 22q11DS and controls, suggesting that the right ALPS index undergoes a modest age-related modulation across development, consistent with physiological maturation rather than a group-specific pathological change. Sex did not emerge as a significant predictor in any model. The summary of all full models is displayed in **Supplementary Table 4**. These findings provide convergent evidence that glymphatic clearance is already compromised in 22q11DS from early childhood (as young as 5 years, the lower bound of our cohort), particularly in the right hemisphere, potentially reflecting early structural or vascular anomalies reported in this population.

### 3.2. Glymphatic Development in Relation to Psychosis Risk within 22q11DS

To evaluate whether glymphatic maturation differs as a function of psychosis liability, we compared ALPS index values across age between 22q11DS individuals with (PPS+) and without (PPS−) clinically significant positive psychotic symptoms. Mixed-effects models revealed no significant group-level difference in ALPS index at the mean age in the bilateral average (p = 0.141; **Figure 3a**) or left hemisphere models (p = 0.955; **Figure 3b**). However, in the right hemisphere, a significant main effect of group was observed (p = 0.025; **Figure 3c**), with PPS+ individuals exhibiting higher ALPS index values than PPS− at the mean age of the sample.

**Figure 3.**
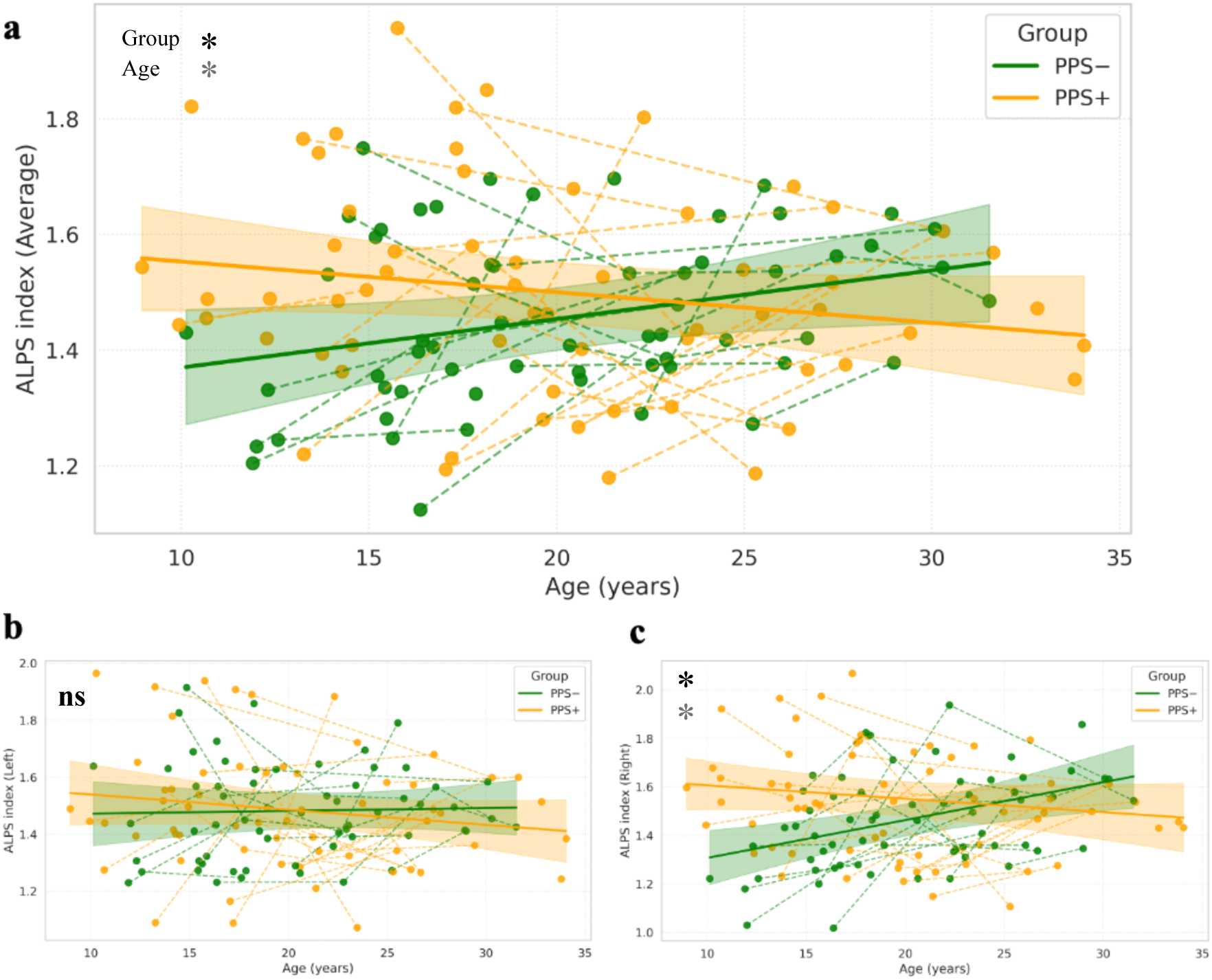
Divergent age-related trajectories of glymphatic efficiency in 22q11DS individuals with and without positive psychotic symptoms. **(a)** Average ALPS index plotted against age in 22q11DS individuals stratified by presence (PPS+, orange) or absence (PPS−, green) of clinically significant positive psychotic symptoms. Each dot represents a scan; dashed lines connect longitudinal data from the same individual. Shaded areas represent 95% confidence intervals of the fitted linear mixed-effects models. Although no significant group-level difference was observed at the mean age (*p* = 0.141), the Group × Age interaction was significant (*p* = 0.009), indicating divergent developmental trajectories: PPS− individuals exhibited increasing glymphatic efficiency with age, while PPS+ individuals showed flat or declining trajectories. **(b)** In the left hemisphere, no significant effects were detected for group (*p* = 0.955), age (*p* = 0.785), or their interaction (*p* = 0.292). **(c)** In the right hemisphere, both the main effect of group (*p* = 0.025) and the Group × Age interaction (*p* = 0.001) were significant. PPS+ individuals showed higher ALPS index values at the mean age compared to PPS−, but failed to exhibit the normative age-related increase in glymphatic efficiency, resulting in a flatter or declining developmental trajectory. All models included age (mean-centered), group, sex, and age-by-group interaction as fixed effects, and subject-level random slopes for age to account for repeated measures. Black asterisks indicate significant group differences, and grey asterisks indicate significant age effects. *p < 0.05, **p < 0.01, ***p < 0.001, ****p < 0.0001; ns = not significant.

Crucially, significant Group × Age interactions were found in both the average and right hemisphere models (average: p = 0.009; right: p = 0.001; **Figure 3a,c**), indicating divergent developmental trajectories between the two subgroups. While PPS− individuals displayed the expected age-related increase in ALPS index, PPS+ individuals showed a markedly blunted or declining trajectory, particularly in the right hemisphere. This pattern suggests that, although PPS+ individuals exhibited elevated ALPS values in early life, their glymphatic function did not follow the age-related increase observed in PPS− individuals.

In the left hemisphere, no significant effects were detected for group (p = 0.955), age (p = 0.785), or their interaction (p = 0.292; **Figure 3b**). Similarly, sex was not a significant predictor in any of the models (Average: p = 0.805; Right: p = 0.504; Left: p = 0.987), indicating that the observed differences in glymphatic trajectories are unlikely to be attributable to sex.

Overall, these findings indicate an alteration in glymphatic development in 22q11DS individuals meeting clinical thresholds for positive psychotic symptoms, which appears more pronounced in the right hemisphere. A full summary of model estimates and significance values is provided in **Supplementary Table 6**.

### 3.3. Glymphatic Function and Excitatory/Inhibitory Imbalance in 22q11DS

Linear regression analyses were performed on 38 scans for the Right and Average ALPS indices, and on 39 scans for the Left ALPS index, after outlier removal as detailed in the *Methods* section. A significant negative association emerged between the ALPS index and the Glx/GABA ratio in the right hippocampus, controlling for age and sex. In particular, the Average ALPS index significantly predicted the Glx/GABA ratio (p = 0.0022), accounting for 25.5% of the variance (R² = 0.255; **Figure 4**).

**Figure 4:**
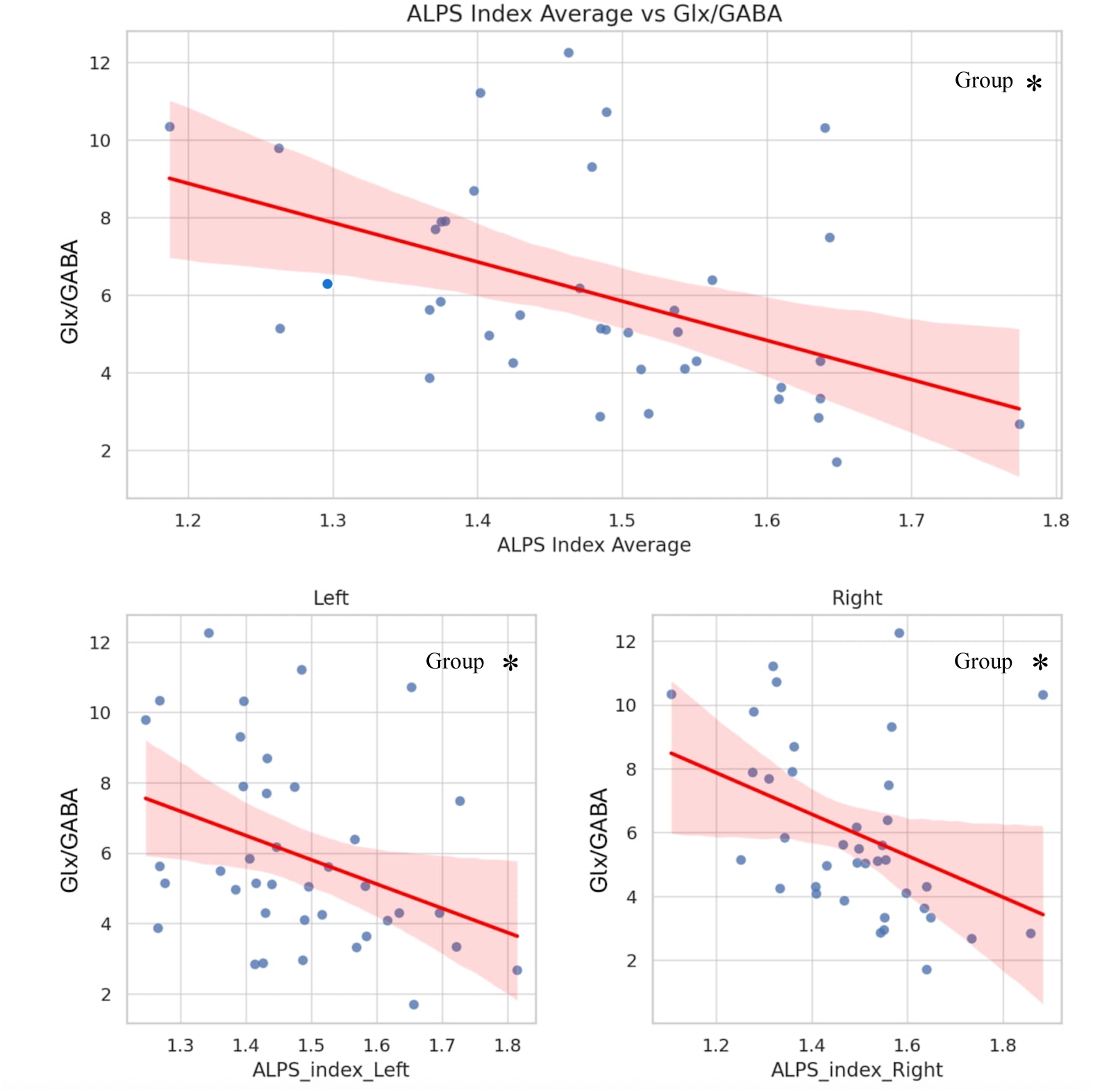
Association between ALPS index and Glx/GABA ratio in 22q11DS. Scatterplots depict the linear association between ALPS index and the Glx/GABA ratio (E/I balance) in the right hippocampus for the bilateral average (top), left hemisphere (bottom left), and right hemisphere (bottom right) ALPS indices. Each dot represents a subject. Red regression lines represent the best-fit line from linear models, with shaded areas indicating the 95% confidence intervals. All models controlled for age and sex. Significant negative associations were observed for all three ALPS metrics, with the strongest effect found for the average index (p = 0.0022). Black asterisks indicate significant group differences, and grey asterisks indicate significant age effects. *p < 0.05, **p < 0.01, ***p < 0.001, ****p < 0.0001; ns = not significant.

When analyzing hemispheres separately, the left ALPS index also significantly predicted the Glx/GABA ratio (p = 0.0181, R² = 0.159), as did the right ALPS index (p = 0.0148, R² = 0.174). These findings indicate that reduced glymphatic efficiency, as indexed by ALPS, is robustly associated with increased excitatory/inhibitory imbalance measured in our cohort in the right hippocampus in 22q11DS, with consistent effects across both hemispheres and the strongest association observed for the bilateral average index.

Full model results are reported in **Supplementary Table 7**.

### 3.4 ALPS Index Comparison between HC and 22q11DS – Automated Method

To rule out potential bias arising from manual ROI placement, we re-analysed the data using an automated pipeline for ALPS index computation, as detailed in the *Methods* section. This approach algorithmically defined ROIs adjacent to the lateral ventricles in each subject’s native space. We then fitted linear mixed-effects models for the Left, Right, and Average ALPS indices, including Group (HC vs. 22q11DS), Age (mean-centred), and Sex as fixed effects, and allowing for a random slope for Age nested within Subject ID to account for within-subject longitudinal variability. A significant main effect of Group was observed for both the Right ALPS index (p = 0.008; **Supplementary Figure 1c**) and the Average ALPS index (p = 0.003; **Supplementary Figure 1a**), reflecting lower ALPS values in individuals with 22q11DS relative to HCs. The Left ALPS index exhibited a non-significant trend in the same direction (p = 0.129; **Supplementary Figure1b**). These findings closely replicated the results obtained using manually defined ROIs, supporting the reliability of ALPS index measurements and reinforcing the robustness of the group differences. Full model results are reported in **Supplementary Table 5.**

## 4. DISCUSSION

### 4.1. Developmental glymphatic inefficiency as a potential early vulnerability marker of psychosis in 22q11DS

The ALPS index, derived from directional diffusivity along perivascular spaces, has been proposed as a non-invasive proxy for assessing glymphatic function (50). To our knowledge, this is the first study to characterize developmental alterations in this metric in the context of psychosis risk. We observed that individuals with 22q11DS already exhibited lower ALPS index values in childhood, potentially indicating early disruption in perivascular fluid dynamics. These findings may reflect the downstream effects of the 22q11.2 deletion on neurovascular (43) and astroglial systems (63), both of which are implicated in the regulation of interstitial clearance (46,64).

One possible contributing factor is the hemizygous deletion of the gene encoding claudin-5 (CLDN5), a tight junction protein expressed in brain microvascular endothelial cells. Prior evidence from animal models suggests that CLDN5 haploinsufficiency may compromise tight junction integrity and increase blood–brain barrier (BBB) permeability, potentially altering perivascular homeostasis (65–67). Increased BBB leakiness could, in turn, affect fluid exchange and directional diffusivity in perivascular spaces, thereby influencing the ALPS index.

Additionally, glymphatic transport is thought to depend on aquaporin-4 (AQP4) water channels located in astrocytic end-feet (2,68). Several genes within the 22q11.2 locus are involved in mitochondrial-dependent processes underlying astrocyte development and function, and astrocytic abnormalities have been reported in animal models of 22q11DS (46,47). Impaired AQP4 localisation or astrocytic polarity may interfere with CSF influx and restrict water diffusivity along perivascular conduits, possibly contributing to the reduced ALPS values observed. While speculative, the combination of BBB dysfunction and astrocytic alterations provides a plausible cellular basis for our results observing an early glymphatic inefficiency in 22q11DS.

Taking advantage of the longitudinal design, we stratified individuals with 22q11DS according to the later emergence of clinically significant positive psychotic symptoms (PPS+ vs PPS−). ALPS trajectories diverged between these groups early in life and prior to the onset of symptoms, suggesting that alterations in perivascular fluid dynamics may precede and potentially contribute to later vulnerability. Interestingly, at baseline, the PPS+ group showed slightly higher ALPS values than the PPS− group. However, over time, PPS− individuals displayed an age-related increase in ALPS, consistent with physiological glymphatic maturation, whereas PPS+ individuals failed to show this developmental progression.

Based on existing literature suggesting that the ALPS index reflects aspects of glymphatic efficiency (50,52), the transient early elevation observed in the PPS+ subgroup could represent a compensatory upregulation in response to early stressors or immune challenges. 22q11DS is associated with broad immune dysregulation, including T-cell deficiency and impaired mucosal immunity (69–72). It is conceivable that only a subset of individuals experiences chronic or recurrent infections, resulting in sustained peripheral and central immune activation. This may elevate the brain’s metabolic and inflammatory load, particularly in perivascular compartments. In this context, an initial increase in ALPS values in PPS+ individuals may reflect a temporary adaptive response to such stressors. However, failure to maintain this compensation over time may signal the collapse of clearance capacity under increasing developmental strain, potentially contributing to downstream neurodevelopmental disruptions and psychosis onset.

### 4.2. Glymphatic clearance and hippocampal excitation/inhibition imbalance

Excitotoxicity has long been proposed as a contributor to psychosis pathophysiology (6,73–75). Excess extracellular glutamate, arising from impaired reuptake, NMDA receptor hypofunction, or altered cortical drive, can lead to oxidative stress, interneuron dysfunction, and structural atrophy (76). Prior studies have linked hippocampal glutamate abnormalities to prodromal symptoms and hippocampal volume loss, in both idiopathic psychosis and 22q11DS (21,22,77).

In this study, we observed a significant inverse correlation between the ALPS index and the Glx/GABA ratio in the right hippocampus. While correlational, these findings are consistent with the hypothesis that impaired glymphatic function may interfere with glutamate clearance, allowing excitatory by-products to accumulate and contributing to local circuit hyperexcitability.

Notably, this association was observed specifically in the right hippocampus, a region known to exhibit high metabolic demand (78) and dense vascularization (79). The hippocampus is also particularly vulnerable to oxidative stress and neuroinflammation during early development (8,75), which may amplify its sensitivity to impaired metabolite clearance. Although hippocampal Glx/GABA ratios may reflect global neurochemical alterations, it is possible that the hippocampus functions as an early sentinel region where such imbalances first become detectable. Future studies employing whole-brain MRS and multimodal imaging will be essential to determine whether this association generalizes beyond the hippocampus.

Taken together, our findings support a developmental framework in which early cellular vulnerabilities in 22q11DS, potentially affecting astrocytic function and BBB integrity, may compromise glymphatic clearance from childhood. This dysfunction appears to persist or worsen in individuals who go on to develop psychotic symptoms (PPS+) and is accompanied by hippocampal excitation/inhibition imbalance. The convergence of impaired interstitial clearance and neurochemical dysregulation may amplify excitotoxic cascades and network instability, potentially contributing to psychosis risk. As illustrated in **Figure 5**, these findings can be conceptualized within a broader mechanistic framework linking the 22q11.2 deletion to increased psychosis risk.

**Figure 5.**
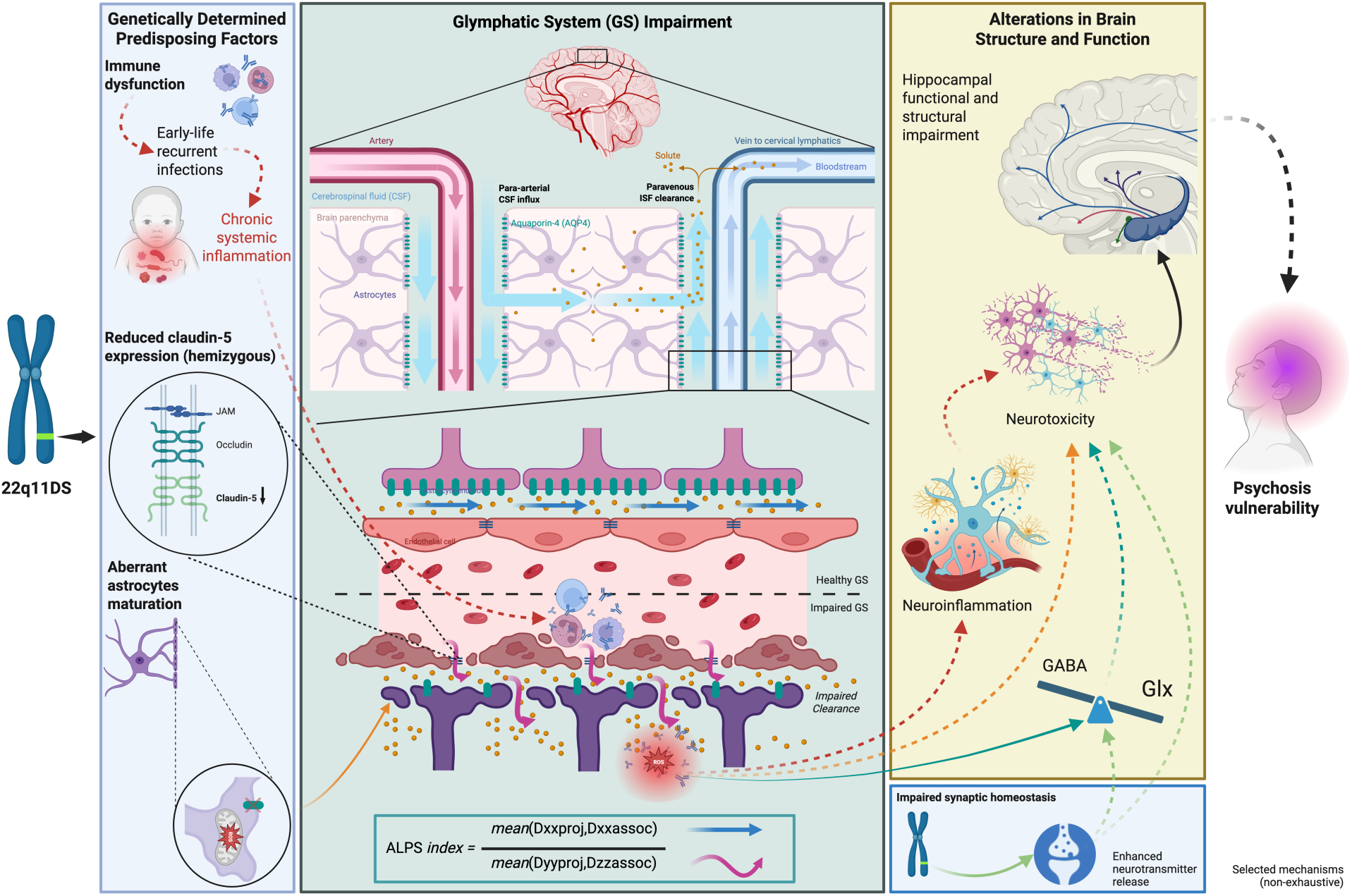
Proposed mechanistic framework linking 22q11.2 deletion to glymphatic dysfunction and psychosis vulnerability. The hemizygous 22q11.2 deletion confers several genetically determined predispositions, including systemic immune dysfunction due to impaired maturation of T lymphocytes and impaired mucosal immunity, leading to increased susceptibility to early-life infections and recurrent systemic inflammation. Additional vulnerabilities include reduced claudin-5 expression, which compromises blood–brain barrier (BBB) integrity, and aberrant astrocytic maturation associated with haploinsufficiency of mitochondrial-related genes within the deleted locus. Mitochondrial deficits increase oxidative stress and impair astrocytic maturation and polarization, thereby disrupting aquaporin-4 (AQP4) localisation at astrocytic end-feet. AQP4 channels are essential for cerebrospinal–interstitial fluid interchange, and their loss of polarity compromises the integrity of the external wall of perivascular spaces, a critical element of glymphatic function. These converging mechanisms are hypothesised to impair glymphatic system (GS) efficiency, leading to reduced perivascular clearance, with the ALPS index serving as a non-invasive proxy of this process. Impaired clearance may promote solute accumulation within the brain parenchyma, contributing to neuroinflammation, excitation/inhibition imbalance (here indexed by Glx/GABA ratios), oxidative stress (ROS increase) and subsequent neurotoxicity. In parallel, haploinsufficiency of presynaptic genes within the deleted locus affect synaptic development and plasticity, compromising homeostatic mechanisms and potentially leading to enhanced neurotransmitter release which might contribute to excitatory–inhibitory (E/I) imbalance, and ultimately neurotoxicity. Due to their high metabolic demand, dense vascularisation, and susceptibility to oxidative stress and excitotoxicity, hippocampal circuits are especially vulnerable and tend to be compromised early in development. Evidence further suggests that hippocampal dysfunction can disrupt dopaminergic circuitry, driving aberrant dopaminergic activity that is closely linked to the emergence of psychotic symptoms. The schematic highlights a non-exhaustive set of mechanisms converging on glymphatic dysfunction and illustrates their interactions across levels, from molecular deficits to clinical phenotype.

While preliminary, this model suggests that glymphatic inefficiency may be a modifiable component of early neurodevelopmental trajectories. Intervening on factors that support interstitial fluid homeostasis, such as sleep, inflammation, or astrocytic function (5,36,80), may offer novel preventive opportunities in populations at high clinical risk.

### 4.3. Limitations and future perspectives

Several limitations should be noted. First, the ALPS index captures only a specific dimension of perivascular diffusivity and may be influenced by confounding factors such as fiber crossings or oedema (52,81). Nevertheless, here we show a robust correlation between ALPS and hippocampal metabolites which supports the biological relevance of ALPS index as a potential proxy for glymphatic clearance.

Second, following the standard published methodology for the calculation of the ALPS index (50), ROI placement was performed manually, which may introduce operator bias. Therefore, we introduce an automated, atlas-based ALPS ROI selection which replicated the main group effects, suggesting the reproducibility of our findings.

Third, group differences in the ALPS index exhibited a right-lateralised pattern, with the most pronounced effects observed in the right hemisphere, although similar trends were present on the left. This hemispheric asymmetry aligns with prior reports of right-lateralised structural and functional anomalies in 22q11DS (82,83) and may reflect increased vulnerability of the right hemisphere to neurodevelopmental perturbations. Notably, right hippocampal atrophy has been implicated in schizophrenia (84) and 22q11DS (85) with altered developmental trajectory of right hippocampal volume in psychotic positive subject emerging on the right side, further supporting the relevance of this asymmetry for psychosis risk. Within our dataset, spectroscopy measures were available only from the right hippocampus and were crosssectional in nature. This limits the interpretation of ALPS–metabolite associations to a unilateral and non-causal level. Future studies employing longitudinal, whole-brain MRS with a wider range of metabolite imaging are warranted to examine ALPS–brain metabolite relationships more comprehensively.

Finally, translational efforts bridging human imaging with mechanistic animal work will be essential to clarify causal pathways. For example, 22q11DS mouse models, such as LgDel, have demonstrated mitochondrial deficits affecting astrocyte development (46,86) and may serve as valuable platforms to investigate how genetic haploinsufficiency perturbs glymphatic architecture and brain clearance function, ultimately impacting circuit dynamics relevant to psychosis. If glymphatic inefficiency contributes to a “second-hit” mechanism in 22q11DS, it may represent a promising target for early intervention focused on restoring physiological homeostasis rather than managing downstream symptoms.

In addition to its relevance for psychosis, glymphatic dysfunction may also contribute to later neurodegenerative vulnerability in 22q11DS. Previous literature has linked impaired glymphatic clearance to reduced elimination of neurotoxic proteins such as α-synuclein, a key pathological hallmark of Parkinson’s disease (PD) (87). Individuals with 22q11DS exhibit a significantly elevated risk for early-onset PD, with recent studies reporting a prevalence of 14% in those over age 50 and a median age at motor onset of 45 years (88). This vulnerability may stem from neurovascular fragility associated with the 22q11.2 deletion, including haploinsufficiency of the CLDN5 gene (66). Given this risk profile, extending longitudinal investigations of glymphatic system function in 22q11DS into the age range when neurodegenerative processes are more likely to manifest would be particularly valuable. Such long-term follow-up could clarify whether glymphatic alterations precede or parallel the onset and progression of parkinsonian symptomatology in this population.

## Data Availability

All data produced in the present study are available upon reasonable request to the corresponding author. Only anonymized datasets will be shared, in order to safeguard participant confidentiality.

## Acknowledgements

We are deeply grateful to all members of the Developmental Imaging and Psychopathology Laboratory for their invaluable support throughout this project. In particular, we warmly thank Caren Latrèche for her outstanding coordination work, and the research assistants collaborating on the project this year and in previous years (Helga Holmquist, Justine Hautekeete and Eva Bertoletti) for their essential contribution to data collection and participant follow-up. We also thank the MRI technicians at Campus Biotech in Geneva for their support during scanning. Finally, we are grateful to all the participants and their families for their trust and continued engagement in our longitudinal study. **Figure 5** is created with BioRender.com.

## Bibliography

1. Chen S, Wang H, Zhang L, Xi Y, Lu Y, Yu K, et al. Glymphatic system: a self-purification circulation in brain. Front Cell Neurosci. 2025 Feb 12;19:1528995.

2. Peng S, Liu J, Liang C, Yang L, Wang G. Aquaporin-4 in glymphatic system, and its implication for central nervous system disorders. Neurobiol Dis. 2023 Apr 1;179:106035.

3. Jessen NA, Munk ASF, Lundgaard I, Nedergaard M. The Glymphatic System: A Beginner’s Guide. Neurochem Res. 2015 Dec 1;40(12):2583–99.

4. Bohr T, Hjorth PG, Holst SC, Hrabětová S, Kiviniemi V, Lilius T, et al. The glymphatic system: Current understanding and modeling. iScience. 2022 Sep 16;25(9):104987.

5. Cai Y, Zhang Y, Leng S, Ma Y, Jiang Q, Wen Q, et al. The relationship between inflammation, impaired glymphatic system, and neurodegenerative disorders: A vicious cycle. Neurobiol Dis. 2024 Mar 1;192:106426.

6. Plitman E, Nakajima S, de la Fuente-Sandoval C, Gerretsen P, Chakravarty MM, Kobylianskii J, et al. Glutamate-mediated excitotoxicity in schizophrenia: A review. Eur Neuropsychopharmacol J Eur Coll Neuropsychopharmacol. 2014 Oct;24(10):1591–605.

7. Murray AJ, Rogers JC, Katshu MZUH, Liddle PF, Upthegrove R. Oxidative Stress and the Pathophysiology and Symptom Profile of Schizophrenia Spectrum Disorders. Front Psychiatry [Internet]. 2021 Jul 22 [cited 2025 Jul 15];12. Available from: https://www.frontiersin.org/journals/psychiatry/articles/10.3389/fpsyt.2021.703452/full

8. Comer AL, Carrier M, Tremblay MÈ, Cruz-Martín A. The Inflamed Brain in Schizophrenia: The Convergence of Genetic and Environmental Risk Factors That Lead to Uncontrolled Neuroinflammation. Front Cell Neurosci [Internet]. 2020 Aug 27 [cited 2025 Jul 15];14. Available from: https://www.frontiersin.org/journals/cellular-neuroscience/articles/10.3389/fncel.2020.00274/full

9. Trépanier MO, Hopperton KE, Mizrahi R, Mechawar N, Bazinet RP. Postmortem evidence of cerebral inflammation in schizophrenia: a systematic review. Mol Psychiatry. 2016 Aug;21(8):1009–26.

10. Lee EE, Hong S, Martin AS, Eyler LT, Jeste DV. Inflammation in Schizophrenia: Cytokine Levels and Their Relationships to Demographic and Clinical Variables. Am J Geriatr Psychiatry Off J Am Assoc Geriatr Psychiatry. 2017 Jan;25(1):50–61.

11. Conen S, Gregory CJ, Hinz R, Smallman R, Corsi-Zuelli F, Deakin B, et al. Neuroinflammation as measured by positron emission tomography in patients with recent onset and established schizophrenia: implications for immune pathogenesis. Mol Psychiatry. 2021 Sep;26(9):5398–406.

12. Warren N, O’Gorman C, Horgan I, Weeratunga M, Halstead S, Moussiopoulou J, et al. Inflammatory cerebrospinal fluid markers in schizophrenia spectrum disorders: A systematic review and meta-analysis of 69 studies with 5710 participants. Schizophr Res. 2024 Apr 1;266:24–31.

13. Steiner J, Mawrin C, Ziegeler A, Bielau H, Ullrich O, Bernstein HG, et al. Distribution of HLA-DR-positive microglia in schizophrenia reflects impaired cerebral lateralization. Acta Neuropathol (Berl). 2006 Sep;112(3):305–16.

14. Gober R, Ardalan M, Shiadeh SMJ, Duque L, Garamszegi SP, Ascona M, et al. Microglia activation in postmortem brains with schizophrenia demonstrates distinct morphological changes between brain regions. Brain Pathol Zurich Switz. 2022 Jan;32(1):e13003.

15. Uliana DL, Lisboa JRF, Gomes FV, Grace AA. The excitatory-inhibitory balance as a target for the development of novel drugs to treat schizophrenia. Biochem Pharmacol. 2024 Oct;228:116298.

16. Lányi O, Koleszár B, Schulze Wenning A, Balogh D, Engh MA, Horváth AA, et al. Excitation/inhibition imbalance in schizophrenia: a meta-analysis of inhibitory and excitatory TMS-EMG paradigms. Schizophrenia. 2024 Jun 15;10(1):56.

17. Merritt K, McGuire P, Egerton A. Relationship between Glutamate Dysfunction and Symptoms and Cognitive Function in Psychosis. Front Psychiatry. 2013 Nov 26;4:151.

18. Rowland LM, Summerfelt A, Wijtenburg SA, Du X, Chiappelli JJ, Krishna N, et al. Frontal Glutamate and γ-Aminobutyric Acid Levels and Their Associations With Mismatch Negativity and Digit Sequencing Task Performance in Schizophrenia. JAMA Psychiatry. 2016 Feb;73(2):166–74.

19. Marsman A, van den Heuvel MP, Klomp DWJ, Kahn RS, Luijten PR, Hulshoff Pol HE. Glutamate in schizophrenia: a focused review and meta-analysis of ^1^H-MRS studies. Schizophr Bull. 2013 Jan;39(1):120–9.

20. Bossong MG, Antoniades M, Azis M, Samson C, Quinn B, Bonoldi I, et al. Association of Hippocampal Glutamate Levels With Adverse Outcomes in Individuals at Clinical High Risk for Psychosis. JAMA Psychiatry. 2019 Feb 1;76(2):199–207.

21. Mancini V, Saleh MG, Delavari F, Bagautdinova J, Eliez S. Excitatory/Inhibitory Imbalance Underlies Hippocampal Atrophy in Individuals With 22q11.2 Deletion Syndrome With Psychotic Symptoms. Biol Psychiatry. 2023 Oct 1;94(7):569–79.

22. Schobel SA, Chaudhury NH, Khan UA, Paniagua B, Styner MA, Asllani I, et al. Imaging patients with psychosis and a mouse model establishes a spreading pattern of hippocampal dysfunction and implicates glutamate as a driver. Neuron. 2013 Apr 10;78(1):81–93.

23. Kraguljac NV, White DM, Reid MA, Lahti AC. Increased Hippocampal Glutamate and Volumetric Deficits in Unmedicated Patients With Schizophrenia. JAMA Psychiatry. 2013 Dec 1;70(12):1294–302.

24. Tu Y, Fang Y, Li G, Xiong F, Gao F. Glymphatic System Dysfunction Underlying Schizophrenia Is Associated With Cognitive Impairment. Schizophr Bull. 2024 Aug 27;50(5):1223–31.

25. Peters ME, Lyketsos CG. The glymphatic system’s role in traumatic brain injury-related neurodegeneration. Mol Psychiatry. 2023 Jul;28(7):2707–15.

26. Gu S, Li Y, Jiang Y, Huang JH, Wang F. Glymphatic Dysfunction Induced Oxidative Stress and Neuro-Inflammation in Major Depression Disorders. Antioxidants. 2022 Nov 20;11(11):2296.

27. Buccellato FR, D’Anca M, Serpente M, Arighi A, Galimberti D. The Role of Glymphatic System in Alzheimer’s and Parkinson’s Disease Pathogenesis. Biomedicines. 2022 Sep 13;10(9):2261.

28. Beschorner N, Nedergaard M. Glymphatic system dysfunction in neurodegenerative diseases. Curr Opin Neurol. 2024 Apr 1;37(2):182–8.

29. Liu H, Chen L, Zhang C, Liu C, Li Y, Cheng L, et al. Glymphatic influx and clearance are perturbed in Huntington’s disease. JCI Insight. 2024 Oct 22;9(20):e172286.

30. Li X, Ruan C, Zibrila AI, Musa M, Wu Y, Zhang Z, et al. Children with autism spectrum disorder present glymphatic system dysfunction evidenced by diffusion tensor imaging along the perivascular space. Medicine (Baltimore). 2022 Dec 2;101(48):e32061.

31. Abdolizadeh A, Torres-Carmona E, Kambari Y, Amaev A, Song J, Ueno F, et al. Evaluation of the Glymphatic System in Schizophrenia Spectrum Disorder Using Proton Magnetic Resonance Spectroscopy Measurement of Brain Macromolecule and Diffusion Tensor Image Analysis Along the Perivascular Space Index. Schizophr Bull. 2024 May 15;sbae060.

32. Wang M, He K, Zhang L, Xu D, Li X, Wang L, et al. Assessment of glymphatic function and white matter integrity in children with autism using multi-parametric MRI and machine learning. Eur Radiol. 2025 Mar 1;35(3):1623–36.

33. Li X, Lin Z, Liu C, Bai R, Wu D, Yang J. Glymphatic Imaging in Pediatrics. J Magn Reson Imaging. 2024;59(5):1523–41.

34. Wong AMC, Siow TY, Cheng YT, Lin ECY, Lin SN, Lin KL, et al. Age-related change of glymphatic function in normative children assessed using diffusion tensor imaging-analysis along the perivascular space. Magn Reson Imaging. 2025 Jul 1;120:110398.

35. Peng T, Lin Y, Xu X, Li J, Liu M, Zhang C, et al. Assessing neonatal brain glymphatic system development using diffusion tensor imaging along the perivascular space and choroid plexus volume. BMC Med Imaging. 2025 Apr 17;25(1):126.

36. Hauglund NL, Andersen M, Tokarska K, Radovanovic T, Kjaerby C, Sørensen FL, et al. Norepinephrine-mediated slow vasomotion drives glymphatic clearance during sleep. Cell. 2025 Feb 6;188(3):606–622.e17.

37. Ma J, Chen M, Liu GH, Gao M, Chen NH, Toh CH, et al. Effects of sleep on the glymphatic functioning and multimodal human brain network affecting memory in older adults. Mol Psychiatry. 2025 May;30(5):1717–29.

38. Castellani G, Ciampoli M, Benedetti A, Ferretti V, Trigilio G, Barcik W, et al. Oxytocin seals the blood-brain barrier, improving 22q11.2 deletion syndrome trajectories. Brain J Neurol. 2025 Jul 15;awaf112.

39. Qin X, Chen J, Zhou T. 22q11.2 deletion syndrome and schizophrenia. Acta Biochim Biophys Sin. 2020 Dec 11;52(11):1181–90.

40. Murphy KC, Jones LA, Owen MJ. High rates of schizophrenia in adults with velo-cardio-facial syndrome. Arch Gen Psychiatry. 1999 Oct;56(10):940–5.

41. Schneider M, Debbané M, Bassett AS, Chow EWC, Fung WLA, van den Bree M, et al. Psychiatric disorders from childhood to adulthood in 22q11.2 deletion syndrome: results from the International Consortium on Brain and Behavior in 22q11.2 Deletion Syndrome. Am J Psychiatry. 2014 Jun;171(6):627–39.

42. Birnbaum R, Weinberger DR. Genetic insights into the neurodevelopmental origins of schizophrenia. Nat Rev Neurosci. 2017 Dec;18(12):727–40.

43. Crockett AM, Ryan SK, Vásquez AH, Canning C, Kanyuch N, Kebir H, et al. Disruption of the blood-brain barrier in 22q11.2 deletion syndrome. Brain J Neurol. 2021 Jun 22;144(5):1351–60.

44. Crockett AM, Kebir H, Anderson SA, Jyonouchi S, Romberg N, Alvarez JI. 22q11.2 Deletion-Associated Blood-Brain Barrier Permeability Potentiates Systemic Capillary Leak Syndrome Neurologic Features. J Clin Immunol. 2024 Apr 5;44(4):87.

45. Li Y, Xia Y, Zhu H, Luu E, Huang G, Sun Y, et al. Investigation of Neurodevelopmental Deficits of 22 q11.2 Deletion Syndrome with a Patient-iPSC-Derived Blood-Brain Barrier Model. Cells. 2021 Sep 28;10(10):2576.

46. Bezzi P, Zehnder T, Laugeray A, Rossi D, Bagni C, Mameli M, et al. Role of Astrocyte Dysfunctions in 22q11 Deletion Syndrome. Biol Psychiatry. 2020 May 1;87(9):S112.

47. de Oliveira Figueiredo EC, Bondiolotti BM, Laugeray A, Bezzi P. Synaptic Plasticity Dysfunctions in the Pathophysiology of 22q11 Deletion Syndrome: Is There a Role for Astrocytes? Int J Mol Sci. 2022 Apr 16;23(8):4412.

48. Eom TY, Han SB, Kim J, Blundon JA, Wang YD, Yu J, et al. Schizophrenia-related microdeletion causes defective ciliary motility and brain ventricle enlargement via microRNA-dependent mechanisms in mice. Nat Commun. 2020 Feb 14;11(1):912.

49. Sinderberry B, Brown S, Hammond P, Stevens AF, Schall U, Murphy DGM, et al. Subtypes in 22q11.2 deletion syndrome associated with behaviour and neurofacial morphology. Res Dev Disabil. 2013 Jan;34(1):116–25.

50. Taoka T, Masutani Y, Kawai H, Nakane T, Matsuoka K, Yasuno F, et al. Evaluation of glymphatic system activity with the diffusion MR technique: diffusion tensor image analysis along the perivascular space (DTI-ALPS) in Alzheimer’s disease cases. Jpn J Radiol. 2017 Apr;35(4):172–8.

51. Taoka T, Ito R, Nakamichi R, Nakane T, Sakai M, Ichikawa K, et al. Diffusion-weighted image analysis along the perivascular space (DWI–ALPS) for evaluating interstitial fluid status: age dependence in normal subjects. Jpn J Radiol. 2022 Sep 1;40(9):894–902.

52. Taoka T, Ito R, Nakamichi R, Nakane T, Kawai H, Naganawa S. Diffusion Tensor Image Analysis ALong the Perivascular Space (DTI-ALPS): Revisiting the Meaning and Significance of the Method. Magn Reson Med Sci. 2024 Apr 2;23(3):268–90.

53. Wood KH, Nenert R, Miften AM, Kent GW, Sleyster M, Memon RA, et al. Diffusion Tensor Imaging-Along the Perivascular-Space Index Is Associated with Disease Progression in Parkinson’s Disease. Mov Disord Off J Mov Disord Soc. 2024 Sep;39(9):1504–13.

54. Schneider M, Armando M, Pontillo M, Vicari S, Debbané M, Schultze-Lutter F, et al. Ultra high risk status and transition to psychosis in 22q11.2 deletion syndrome. World Psychiatry. 2016 Oct;15(3):259–65.

55. Delavari F, Rafi H, Sandini C, Murray RJ, Latrèche C, Van De Ville D, et al. Amygdala subdivisions exhibit aberrant whole-brain functional connectivity in relation to stress intolerance and psychotic symptoms in 22q11.2DS. Transl Psychiatry. 2023 May 4;13(1):145.

56. Mancini V, Zöller D, Schneider M, Schaer M, Eliez S. Abnormal Development and Dysconnectivity of Distinct Thalamic Nuclei in Patients With 22q11.2 Deletion Syndrome Experiencing Auditory Hallucinations. Biol Psychiatry Cogn Neurosci Neuroimaging. 2020 Sep 1;5(9):875–90.

57. Tang SX, Yi JJ, Moore TM, Calkins ME, Kohler CG, Whinna DA, et al. Subthreshold Psychotic Symptoms in 22q11.2 Deletion Syndrome. J Am Acad Child Adolesc Psychiatry. 2014 Sep;53(9):991–1000.e2.

58. Weisman O, Guri Y, Gur RE, McDonald-McGinn DM, Calkins ME, Tang SX, et al. Subthreshold Psychosis in 22q11.2 Deletion Syndrome: Multisite Naturalistic Study. Schizophr Bull. 2017 Sep 1;43(5):1079–89.

59. Saito Y, Hayakawa Y, Kamagata K, Kikuta J, Mita T, Andica C, et al. Glymphatic system impairment in sleep disruption: diffusion tensor image analysis along the perivascular space (DTI-ALPS). Jpn J Radiol. 2023 Dec;41(12):1335–43.

60. Tatekawa H, Matsushita S, Ueda D, Takita H, Horiuchi D, Atsukawa N, et al. Improved reproducibility of diffusion tensor image analysis along the perivascular space (DTI-ALPS) index: an analysis of reorientation technique of the OASIS-3 dataset. Jpn J Radiol. 2023 Apr 1;41(4):393–400.

61. Wenneberg C, Nordentoft M, Rostrup E, Glenthøj LB, Bojesen KB, Fagerlund B, et al. Cerebral Glutamate and Gamma-Aminobutyric Acid Levels in Individuals at Ultra-high Risk for Psychosis and the Association With Clinical Symptoms and Cognition. Biol Psychiatry Cogn Neurosci Neuroimaging. 2020 Jun;5(6):569–79.

62. Saleh MG, Papantoni A, Mikkelsen M, Hui SCN, Oeltzschner G, Puts NA, et al. Effect of Age on GABA+ and Glutathione in a Pediatric Sample. AJNR Am J Neuroradiol. 2020 Jun;41(6):1099–104.

63. Tegtmeyer M, Liyanage D, Han Y, Hebert KB, Pei R, Way GP, et al. Combining phenomics with transcriptomics reveals cell-type-specific morphological and molecular signatures of the 22q11.2 deletion. Nat Commun. 2025 Jul 9;16(1):6332.

64. Abbott NJ, Pizzo ME, Preston JE, Janigro D, Thorne RG. The role of brain barriers in fluid movement in the CNS: is there a ‘glymphatic’ system? Acta Neuropathol (Berl). 2018 Mar 1;135(3):387–407.

65. Sun ZY, Wei J, Xie L, Shen Y, Liu SZ, Ju GZ, et al. The CLDN5 locus may be involved in the vulnerability to schizophrenia. Eur Psychiatry. 2004 Sep;19(6):354–7.

66. Hashimoto Y, Greene C, Munnich A, Campbell M. The CLDN5 gene at the blood-brain barrier in health and disease. Fluids Barriers CNS. 2023 Mar 28;20:22.

67. Arinami T. Analyses of the associations between the genes of 22q11 deletion syndrome and schizophrenia. J Hum Genet. 2006 Dec;51(12):1037–45.

68. Iliff JJ, Wang M, Liao Y, Plogg BA, Peng W, Gundersen GA, et al. A paravascular pathway facilitates CSF flow through the brain parenchyma and the clearance of interstitial solutes, including amyloid β. Sci Transl Med. 2012 Aug 15;4(147):147ra111.

69. Morsheimer M, Brown Whitehorn TF, Heimall J, Sullivan KE. The immune deficiency of chromosome 22q11.2 deletion syndrome. Am J Med Genet A. 2017 Sep;173(9):2366–72.

70. McDonald-McGinn DM, Sullivan KE, Marino B, Philip N, Swillen A, Vorstman JAS, et al. 22q11.2 deletion syndrome. Nat Rev Dis Primer. 2015 Nov 19;1(1):15071.

71. Jawad AF, McDonald-Mcginn DM, Zackai E, Sullivan KE. Immunologic features of chromosome 22q11.2 deletion syndrome (DiGeorge syndrome/velocardiofacial syndrome). J Pediatr. 2001 Nov;139(5):715–23.

72. Gennery AR. Immunological aspects of 22q11.2 deletion syndrome. Cell Mol Life Sci CMLS. 2011 Oct 9;69(1):17–27.

73. Olney JW, Farber NB. Glutamate receptor dysfunction and schizophrenia. Arch Gen Psychiatry. 1995 Dec;52(12):998–1007.

74. Moghaddam B, Javitt D. From Revolution to Evolution: The Glutamate Hypothesis of Schizophrenia and its Implication for Treatment. Neuropsychopharmacology. 2012 Jan;37(1):4–15.

75. Steullet P, Cabungcal JH, Monin A, Dwir D, O’Donnell P, Cuenod M, et al. Redox dysregulation, neuroinflammation, and NMDA receptor hypofunction: A “central hub” in schizophrenia pathophysiology? Schizophr Res. 2016 Sep;176(1):41–51.

76. Verma M, Lizama BN, Chu CT. Excitotoxicity, calcium and mitochondria: a triad in synaptic neurodegeneration. Transl Neurodegener. 2022 Jan 25;11(1):3.

77. Basavaraju R, Guo J, Small SA, Lieberman JA, Girgis RR, Provenzano FA. Hippocampal Glutamate and Positive Symptom Severity in Clinical High Risk for Psychosis. JAMA Psychiatry. 2022 Feb 1;79(2):178–9.

78. Manganas LN, Zhang X, Li Y, Hazel RD, Smith SD, Wagshul ME, et al. Magnetic resonance spectroscopy identifies neural progenitor cells in the live human brain. Science. 2007 Nov 9;318(5852):980–5.

79. Johnson AC. The Hippocampal Vascular Supply and Its Role in Vascular Cognitive Impairment. Stroke. 2023 Mar;54(3):673–85.

80. Das N, Dhamija R, Sarkar S. The role of astrocytes in the glymphatic network: a narrative review. Metab Brain Dis. 2024 Mar 1;39(3):453–65.

81. Ringstad G. Glymphatic imaging: a critical look at the DTI-ALPS index. Neuroradiology. 2024 Feb 1;66(2):157–60.

82. Scariati E, Schaer M, Richiardi J, Schneider M, Debbané M, Van De Ville D, et al. Identifying 22q11.2 Deletion Syndrome and Psychosis Using Resting-State Connectivity Patterns. Brain Topogr. 2014 Nov 1;27(6):808–21.

83. Scariati E, Schaer M, Karahanoglu I, Schneider M, Richiardi J, Debbané M, et al. Largescale functional network reorganization in 22q11.2 deletion syndrome revealed by modularity analysis. Cortex J Devoted Study Nerv Syst Behav. 2016 Sep;82:86–99.

84. Lee K, Lee YM, Park JM, Lee BD, Moon E, Jeong HJ, et al. Right hippocampus atrophy is independently associated with Alzheimer’s disease with psychosis. Psychogeriatr Off J Jpn Psychogeriatr Soc. 2019 Mar;19(2):105–10.

85. Mancini V, Sandini C, Padula MC, Zöller D, Schneider M, Schaer M, et al. Positive psychotic symptoms are associated with divergent developmental trajectories of hippocampal volume during late adolescence in patients with 22q11DS. Mol Psychiatry. 2020 Nov;25(11):2844–59.

86. Fernandez A, Meechan DW, Karpinski BA, Paronett EM, Bryan CA, Rutz HL, et al. Mitochondrial Dysfunction Leads to Cortical Under-Connectivity and Cognitive Impairment. Neuron. 2019 Jun 19;102(6):1127–1142.e3.

87. Yue Y, Zhang X, Lv W, Lai HY, Shen T. Interplay between the glymphatic system and neurotoxic proteins in Parkinson’s disease and related disorders: current knowledge and future directions. Neural Regen Res. 2023 Dec 15;19(9):1973–80.

88. von Scheibler ENMM, Swillen A, Repetto GM, Reyes NGD, Lang AE, Marras C, et al. Prevalence of Parkinson’s Disease in 22q11.2 Deletion Syndrome: A Multicenter Study. Mov Disord Clin Pract. 2025 Jun;12(6):817–22.

