## Supplementary Material (Methods, Tables and Figures) for "Developmental Glymphatic Dysfunction Underlies Excitation/Inhibition Imbalance and Psychosis Vulnerability in 22q11.2 Deletion Syndrome"

**TABLE OF CONTENTS**


1. **SUPPLEMENTARY METHODS**
2. **SUPPLEMENTARY TABLES AND SUPPLEMENTARY TABLES LEGENDS**
3. **SUPPLEMENTARY FIGURES**
4. **SUPPLEMENTARY REFERENCES**


### 1. SUPPLEMENTARY METHODS

*Data Acquisition*

Brain MRI, including Diffusion-weighted images and $^{1}$H-MRS were acquired using a single-shot echo-planar imaging (EPI) sequence on a 3 Tesla Siemens Trio MRI scanner (Siemens Healthineers, Erlangen, Germany). The acquisition protocol included 30 diffusion directions with a b-value of 1000 s/mm$^2$. Imaging parameters were as follows: repetition time (TR) = 8300–8800 ms, echo time (TE) = 84 ms, flip angle = 90°–180°, matrix size = 128 × 128, field of view = 25.6 cm, slice thickness = 2 mm, and isotropic voxel resolution of 2.0 × 2.0 × 2.0 mm$^3$. Sixty-four axial slices were acquired with anterior-to-posterior phase encoding, and GRAPPA parallel imaging was used with an acceleration factor of 2. The T1 images were used to guide $^{1}$H-MRS voxel positioning in the right hippocampus following same protocol as previously published (1).

Participants were scanned using the Mescher-Garwood point-resolved spectroscopy (MEGA-PRESS) Siemens prototype sequence (2) for the measurement of GABA with macromolecules and homocarnosine (GABA+) at 3.0 ppm with the following parameters: repetition time/echo time 1500/68 ms; spectral width: 2000 Hz; editing pulse offset (ON/OFF) 1.9/7.5 ppm; bandwidth of the editing pulse specified on the scanner 60 Hz; 2048 data points; and voxel sizes and number of transients 15 × 35 × 40 $mm^3$ and 196 for ACC, 20 × 30 × 40 $mm^3$ and 300 for right hippocampus, and 30 × 30 × 30 $mm^3$ and 240 for STC. Before every MRS acquisition, we performed shimming using the vendor-specific shim tool. We further optimized the shim values by manually adjusting the linear shim terms. Finally, unsuppressed water acquisitions were performed with the same region-specific parameters, except 16 transients for concentration quantification. All participants were scanned using the same protocol across sessions. Whenever possible, foam padding was used to minimize head motion, and participants were instructed to remain still during the acquisition.

*Diffusion Weighted-Imaging Pre-Processing*

Diffusion-weighted imaging (DWI) data were preprocessed using a standardized pipeline based on MRtrix3 and FSL (v6.0.7.12), following established practices in ALPS-based studies (3) (Taoka et al., 2017). Preprocessing steps included denoising, removal of Gibbs ringing artefacts, correction for motion and eddy current distortions, bias field correction, and brain masking. Diffusion tensors were then fitted using FSL's dtifit, and scalar maps including fractional anisotropy (FA) and mean diffusivity (MD) were computed. FA maps were reoriented to the JHU-ICBM-FA-1mm template using a 6-degree-of-freedom transformation to ensure that the Dxx, Dyy, and Dzz components were aligned with respect to the X, Y, and Z axes, thereby accounting for potential head movement effects that could influence ROI placement. All ALPS index computations were performed in native diffusion space. All preprocessed scans underwent thorough visual quality control to exclude motion artefacts, misalignment, and signal dropout. Details of the preprocessing code and parameters are available on demand.

*ALPS Index Computation*

Following the main preprocessing steps, tensor-derived diffusion metrics were processed using a custom FSL-based pipeline (v6.0.7.12) to extract directional components required for ALPS index computation. In this study, we employed two complementary ROI placement strategies: (1) a manual method, performed in native diffusion space (2) an atlas-based automated registration method to enable reproducible ROI placement adapted to individual anatomy.

For each subject, the three diagonal elements of the diffusion tensor (Dxx, Dyy, Dzz) were isolated using FSL's fslroi, enabling direction-specific analysis of water diffusivity along the right–left (x), anterior–posterior (y), and inferior–superior (z) axes. These components were later used to compute the ALPS index based on established methodology. Additionally, a directional fractional anisotropy (FA) colormap was generated for each scan by multiplying the principal eigenvector (V1) map by the corresponding FA map using FSL's fslmaths. This visualization facilitated quality control and enabled the assessment of local diffusion orientation patterns.

The ALPS index was computed following the methodology described by Taoka et al. (3–5).

*Manual ROI placement method*

Spherical regions of interest (ROIs) with a diameter of 4 mm were manually placed bilaterally at the level of the lateral ventricles. ROIs were placed in white matter regions corresponding to projection and association fibers, whose predominant orientations (z- and y-axes, respectively) were approximated based on local diffusion directions within native space as shown in **Figure 1a**. Placement was performed using FSLeyes, guided by directionally encoded color (DEC) maps and the tensor component images (Dxx, Dyy, Dzz) to ensure anatomical precision.

ROIs were carefully positioned adjacent to medullary veins, which course predominantly along the right–left (x-axis) direction in periventricular regions and are thought to align with perivascular spaces. To enhance measurement specificity, regions where association fibers intersected with subcortical tracts oriented in the x-direction were systematically avoided, in accordance with published recommendations (5).

From each ROI, directional diffusivity values were extracted from the tensor component maps. The ALPS index was then computed using the formula:

**ALPS index = mean(Dx_proj, Dx_assoc) / mean(Dy_proj, Dz_assoc)**

In this equation, the numerator captures diffusivity along the x-axis (presumed to follow perivascular spaces), while the denominator reflects diffusion orthogonal to PVSs, within structured white matter tracts. A higher ALPS index suggests greater fluid mobility along perivascular pathways and is interpreted as an indicator of more efficient glymphatic function.

The ALPS index was calculated separately for the left and right hemispheres in each scan. A bilateral mean was derived by averaging the two hemispheric values.

*Automated ROI Placement method as replication*

To enable automated subject-specific ROI placement, we used a symmetric diffeomorphic registration approach via ANTs. First, the MNI FA template (JHU-ICBM-FA-2mm) was aligned to each subject's native FA map using antsRegistrationSyNQuick.sh with the -t s option. The resulting affine (0GenericAffine.mat) and nonlinear warp (1Warp.nii.gz) transforms were applied to each ROI using antsApplyTransforms, producing individualized ROIs in diffusion space. ROIs were placed bilaterally in the superior corona radiata (SCR) and superior longitudinal fasciculus (SLF), based on established anatomical definitions. After transformation, diffusivity values were extracted from the Dxx, Dyy, and Dzz maps within each ROI. The ALPS index was computed per hemisphere using the formula provided in the main text.

*Supplementary Methods: Statistical Analysis Details*

Statistical analyses were designed to account for the nested structure of the longitudinal dataset, which included repeated DTI measurements acquired across development in both individuals with 22q11DS and HCs. As described above, a total of 258 scans were retained. All scans passed both quality control and preprocessing steps and were included in the final analysis. The distribution of longitudinal scans per participant and inter-scan intervals is reported in **Supplementary Table 2**.

ALPS indices were computed separately for the left and right hemispheres, with outliers removed using interquartile range (IQR ±1.5) filtering applied independently to each side and separately within each group (HC and 22q11DS). Bilateral average ALPS values were then calculated using only scans that retained valid indices in both hemispheres. Detailed counts of retained scans after outlier removal are provided for both manual and automated method in **Supplementary Table 3.**

Psychosis status was assessed using the Structured Interview for Psychosis-Risk Syndromes (SIPS). Of the 85 individuals with 22q11DS, 78 had at least one valid SIPS assessment and were therefore included in the symptom-stratified analyses. Seven individuals were excluded due to missing or invalid SIPS data. Among the included participants, 38 met criteria for moderate-to-severe positive psychotic symptoms (SIPS P1–P5 $\geq 3$) at any timepoint and were classified as psychosis-positive (PPS+), while the remaining 40 were considered psychosis-negative (PPS−). A total of 123 scans from these 78 individuals were retained for the PPS+ vs PPS− analyses (PPS+ = 62 scans, PPS− = 61 scans). As in the main group-level analysis, outliers were removed using the interquartile range (IQR ±1.5) method, applied separately for each hemisphere and independently within each subgroup.

All statistical analyses used the ALPS indices as dependent variables. To account for repeated measures and inter-individual developmental variability, linear mixed-effects models were implemented. These models included fixed effects for age, sex, and diagnostic group, and a random slope for age at the subject level to capture within-subject variability over time. To assess developmental trajectories, we incorporated interaction terms between age and diagnostic group. A linear age term was employed for both the 22q11DS vs. HC and PPS+ vs. PPS− comparisons, providing a consistent and interpretable framework for group-by-age interaction effects. The decision to use a linear specification was based on preliminary analyses, which showed no strong evidence for nonlinear age effects and favored model stability. For interpretability, age was mean-centered prior to analysis. This centering approach allows the main group effect to reflect differences in the ALPS index at the average age of the sample, rather than at an arbitrary zero point outside the observed age range. The final model thus included fixed effects for mean-centered age, diagnostic group (coded as 0 = control or PPS−, 1 = 22q11DS or PPS+), their interaction, and sex, with a subject-level random slope for age to account for repeated measurements.

The same statistical modelling approach was applied to ALPS indices derived from both the manual and automated ROI methods, ensuring consistency in the evaluation of developmental trajectories and group effects.

All statistical analyses were conducted in Python (using `statsmodels` and `pingouin`), and all models used fixed effects for group, age, and sex, with subject ID modelled as a random intercept and random slope to account for repeated measures. Statistical significance was set at $p < 0.05$.

*MRS Processing and Analysis*

All spectral data were processed using Gannet (version 3.1.5) (6). After frequency and phase correction of transients, data were combined to generate GABA-edited spectra. The GABA+, glutamate+glutamine (Glx), and unsuppressed water signals were modeled to calculate GABA and Glx concentrations in international units. T1-weighted images were segmented using SPM12 (7) to calculate gray matter, white matter, and cerebrospinal fluid voxel tissue fractions in the $^1$H-MRS voxels. Subsequently, metabolite concentrations were corrected for partial volume effects using the tissue fractions and relaxation effects with the Gasparovic method (8). Data quality was assessed using the creatine signal at 3 ppm to estimate main magnetic field ($B_0$) drift (before frequency and phase correction) and using the unsuppressed water signal linewidth at full width at half maximum. Software that uses linear combination modeling determines Cramér-Rao lower bounds to express the uncertainty of the fitted signal amplitude. However, Gannet does not provide Cramér-Rao lower bounds but determines fitting quality (called fit error) through the standard deviation of fitting residuals normalized to the height of the modeled peak. The GABA and Glx fit errors generated by Gannet were used for assessing modeling errors in every region. As in previous papers (1,9,10), data with fit errors exceeding 2 standard deviations above the mean of all the data (15%) were excluded from further analyses.

The Glx/GABA ratio was computed and served as a proxy for excitatory/inhibitory (E/I) balance as performed in previous studies (11).

In a subset of 39 participants with 22q11DS (25 males, 14 females; mean age = 21.6 ± 6.9 years), both high-quality spectroscopy and diffusion data were available.

Linear regression models tested the association between ALPS index average (predictor) and Glx/GABA ratio (outcome), controlling for age and sex. Additional regressions using left and right ALPS values separately were performed to assess hemispheric specificity.

Outlier removal was performed using the interquartile range (IQR ±1.5) method, consistent with the statistical procedures applied throughout the study. Outliers were assessed independently for each ALPS index (left and right) as well as for the Glx/GABA ratio. The mean ALPS index was computed only when both left and right values were within non-outlier bounds. This procedure resulted in the exclusion of a single scan based on a low right ALPS index. No outliers were identified for the Glx/GABA ratio. Consequently, 38 scans were retained for the analyses involving ALPS right and ALPS average, while all 39 scans were included in the model using ALPS left. All data were from unique individuals (i.e., no longitudinal scans), and all models included age and sex as covariates.

### 2. SUPPLEMENTARY TABLES AND SUPPLEMENTARY TABLES LEGENDS

**Supplementary Table 1. Overview of Sample Characteristics and Scan Distribution.**

| Group | N Participants | N Scans | Mean Age (± SD) | Sex (M/F) |
|---|---|---|---|---|
| 22q11DS | 85 | 143 | 18.92 ± 6.36 | 43 / 42 |
| Healthy Controls (HC) | 83 | 115 | 16.28 ± 6.42 | 40 / 43 |
| **Total** | **168** | **258** | — | — |

**Supplementary Table 2. Retained ALPS Index Values After Outlier Removal for Manual and Automated Calculation Methods. Values reflect the number of scans retained for each index (Left, Right, Avg) after removing outliers. Average values were retained only if the scan passed outlier thresholds on both the left and right indices.**

| Method | N Before Outlier Removal | Left Retained (No Outliers) | Right Retained (No Outliers) | Avg Retained (No Outliers) |
|---|---|---|---|---|
| Manual | 258 | 250 | 256 | 249 |
| Automated | 258 | 248 | 253 | 246 |

**Supplementary table 3. SIPS Assessment and Psychosis Stratification (22q11DS only).**

| **Assessment Detail** | **n Participants** | **N Scan** |
|---|---|---|
| With valid SIPS data | 78 | 123 |
| Psychosis-positive (SIPS ≥ 3) | 38 | 62 |
| Psychosis-negative | 40 | 61 |
| Without SIPS (e.g., age < 6) | 7 | |

**Supplementary Table 4. Fixed effects from linear mixed-effects models comparing ALPS index values between 22q11DS individuals and HCs.**

Separate models were run for the bilateral average (**Supplementary Table 4a**), left (**Supplementary Table 4b**) and right (**Supplementary Table 4c**) hemisphere indices. Fixed effects included age (mean-centered), group (0 = control, 1 = 22q11DS), sex, and their interaction. Bold *p*-values indicate statistical significance ($p < 0.05$).

### Supplementary Table 4a

| | avg_ind | | |
|---|---|---|---|
| *Predictors* | *Estimates* | *CI* | *p* |
| (Intercept) | 1.54 | 1.50 – 1.58 | **<0.001** |
| Age centered | 0.00 | -0.00 – 0.01 | 0.126 |
| VCFS Status [1] | -0.05 | -0.10 – -0.01 | **0.017** |
| Sex [1] | -0.00 | -0.05 – 0.04 | 0.906 |
| Age centered × VCFS Status [1] | -0.00 | -0.01 – 0.00 | 0.235 |

### Supplementary Table 4b

| | ALPS_index_Left | | |
|---|---|---|---|
| *Predictors* | *Estimates* | *CI* | *p* |
| (Intercept) | 1.51 | 1.47 – 1.56 | **<0.001** |
| Age centered | 0.00 | -0.00 – 0.01 | 0.654 |
| VCFS Status [1] | -0.03 | -0.08 – 0.02 | 0.253 |
| Sex [1] | 0.01 | -0.04 – 0.06 | 0.630 |
| Age centered × VCFS Status [1] | -0.00 | -0.01 – 0.00 | 0.235 |

**Supplementary Table 4c**

| | ALPS_index_Right | | |
|---|---|---|---|
| *Predictors* | *Estimates* | *CI* | *p* |
| (Intercept) | 1.56 | 1.51 – 1.61 | **<0.001** |
| Age centered | 0.01 | 0.00 – 0.01 | **0.037** |
| VCFS Status [1] | -0.07 | -0.13 – -0.01 | **0.022** |
| Sex [1] | -0.00 | -0.06 – 0.06 | 0.890 |
| Age centered × VCFS Status [1] | -0.00 | -0.01 – 0.01 | 0.508 |

**Supplementary Table 5. Fixed effects from linear mixed-effects models comparing ALPS index values between 22q11DS individuals and HCs using the automated ROI method.**

**Supplementary Table 5a: ALPS Index automated - Average**

**Fixed Effects Coefficients:**

| Term | Coef. | Std.Err. | z | p | [0.025, 0.975] |
|---|---|---|---|---|---|
| Intercept | 1.441 | 0.016 | 90.62 | 0.000 | [1.410, 1.472] |
| Sex[T.Male] | 0.023 | 0.018 | 1.27 | 0.202 | [-0.012, 0.057] |
| VCFS_Status | -0.053 | 0.018 | -2.92 | **0.003** | [-0.089, -0.017] |
| Age_centered | -0.002 | 0.002 | -1.10 | 0.271 | [-0.006, 0.002] |
| VCFS_Status x Age_centered | -0.001 | 0.003 | -0.49 | 0.622 | [-0.007, 0.004] |

**Supplementary Table 5b: ALPS Index automated - Left**

**Fixed Effects Coefficients:**

| Term | Coef. | Std.Err. | z | p | [0.025, 0.975] |
|---|---|---|---|---|---|
| Intercept | 1.449 | 0.021 | 69.60 | 0.000 | [1.409, 1.490] |
| Sex[T.Male] | 0.065 | 0.020 | 3.26 | **0.001** | [0.026, 0.104] |
| VCFS_Status | -0.031 | 0.021 | -1.52 | 0.129 | [-0.072, 0.009] |
| Age_centered | 0.020 | 160438.923 | 0.00 | 1.000 | [-314454.491, 314454.531] |
| VCFS_Status x Age_centered | 0.013 | 223018.680 | 0.00 | 1.000 | [-437108.567, 437108.593] |

**Supplementary Table 5c: ALPS Index automated - Right:**

**Fixed Effects Coefficients:**

| Term | Coef. | Std.Err. | z | p | [0.025, 0.975] |
|---|---|---|---|---|---|
| Intercept | 1.436 | 0.024 | 59.68 | 0.000 | [1.389, 1.483] |
| Sex[T.Male] | -0.025 | 0.023 | -1.09 | 0.276 | [-0.069, 0.020] |
| VCFS_Status | -0.063 | 0.024 | -2.64 | **0.008** | [-0.110, -0.016] |
| Age_centered | 0.065 | 179175.052 | 0.00 | 1.000 | [-351176.584, 351176.714] |
| VCFS_Status x Age_centered | -0.013 | nan | nan | nan | [nan, nan] |

**Supplementary Table 6. Fixed effects from linear mixed-effects models predicting ALPS index values as a function of age, group (0 = PPS−, 1 = PPS+), sex, and their interaction.** Separate models were fitted for the bilateral average, right, and left hemisphere indices. Age was mean-centered. Bold *p*-values indicate statistical significance ($p < 0.05$).

### Supplementary Table 6a

| | ALPS_index_Avg | | |
|---|---|---|---|
| *Predictors* | *Estimates* | *CI* | *p* |
| (Intercept) | 1.45 | 1.40 – 1.51 | **<0.001** |
| Age centered | 0.01 | 0.00 – 0.02 | **0.039** |
| Group [1] | 0.05 | -0.02 – 0.11 | 0.141 |
| Sex [1] | 0.01 | -0.05 – 0.07 | 0.805 |
| Age centered × Group [1] | -0.01 | -0.02 – -0.00 | **0.009** |

### Supplementary Table 6b

| | ALPS_index_Left | | |
|---|---|---|---|
| *Predictors* | *Estimates* | *CI* | *p* |
| (Intercept) | 1.48 | 1.42 – 1.54 | **<0.001** |
| Age centered | 0.00 | -0.01 – 0.01 | 0.785 |
| Group [1] | 0.00 | -0.07 – 0.07 | 0.955 |
| Sex [1] | -0.00 | -0.07 – 0.07 | 0.987 |
| Age centered × Group [1] | -0.01 | -0.02 – 0.01 | 0.292 |

**Supplementary Table 6c**

| | ALPS_index_Right | | |
|---|---|---|---|
| *Predictors* | *Estimates* | *CI* | *p* |
| (Intercept) | 1.44 | 1.37 – 1.51 | **<0.001** |
| Age centered | 0.02 | 0.01 – 0.03 | **0.001** |
| Group [1] | 0.09 | 0.01 – 0.17 | **0.025** |
| Sex [1] | 0.03 | -0.05 – 0.11 | 0.504 |
| Age centered × Group [1] | -0.02 | -0.03 – -0.01 | **0.001** |

**Supplementary table 7: Results from linear regression models testing the association between ALPS index and Glx/GABA ratio.**

Each model included one ALPS index (left, right, or average) as the predictor and Glx/GABA ratio (measured in the right hippocampus) as the dependent variable. All models controlled for age and sex. Table reports beta coefficients, p-values for each predictor, and the model $R^2$. Significant associations between ALPS and Glx/GABA ratio were found in all models, indicating a robust inverse relationship between glymphatic function and excitatory/inhibitory imbalance in 22q11DS.

| **ALPS Predictor** | β (ALPS) | p (ALPS) | p (age) | p (Sex) | $R^2$ |
|---|---|---|---|---|---|
| ALPS_index_Left | -7.662 | **0.0181** | 0.3338 | 0.8502 | 0.159 |
| ALPS_index_Right | -6.765 | **0.0148** | 0.4562 | 0.7327 | 0.174 |
| ALPS_index_Average | -11.015 | **0.0022** | 0.2482 | 0.7912 | 0.255 |

### 3. SUPPLEMENTARY FIGURES

**Supplementary Figure 1. Age-related trajectories of ALPS index based on automated ROI placement.**

**(a)** Average ALPS index plotted against age in individuals with 22q11.2 deletion syndrome (22q11DS; red) and healthy controls (HC; blue), using values derived from the automated ROI extraction pipeline. Each dot represents a scan; dashed lines connect longitudinal scans from the same subject. Shaded areas represent 95% confidence intervals of the fitted linear mixed-effects models. A significant group difference was observed, with lower average ALPS values in the 22q11DS group ($p = 0.003$).

**(b)** Left hemisphere ALPS index showed a non-significant trend toward reduced values in 22q11DS compared to HC ($p = 0.129$). Sex was a significant predictor, with higher ALPS index in males compared to females ($p = 0.001$).

**(c)** Right hemisphere ALPS index revealed a significant group difference, with lower values in the 22q11DS group ($p = 0.008$).

All models included group, age (mean-centered), sex, and subject-level random slopes for age to account for repeated measures. Black asterisks indicate significant group differences, and grey asterisks indicate significant age effects. *$p < 0.05$, **$p < 0.01$, ***$p < 0.001$, ****$p < 0.0001$; ns = not significant.

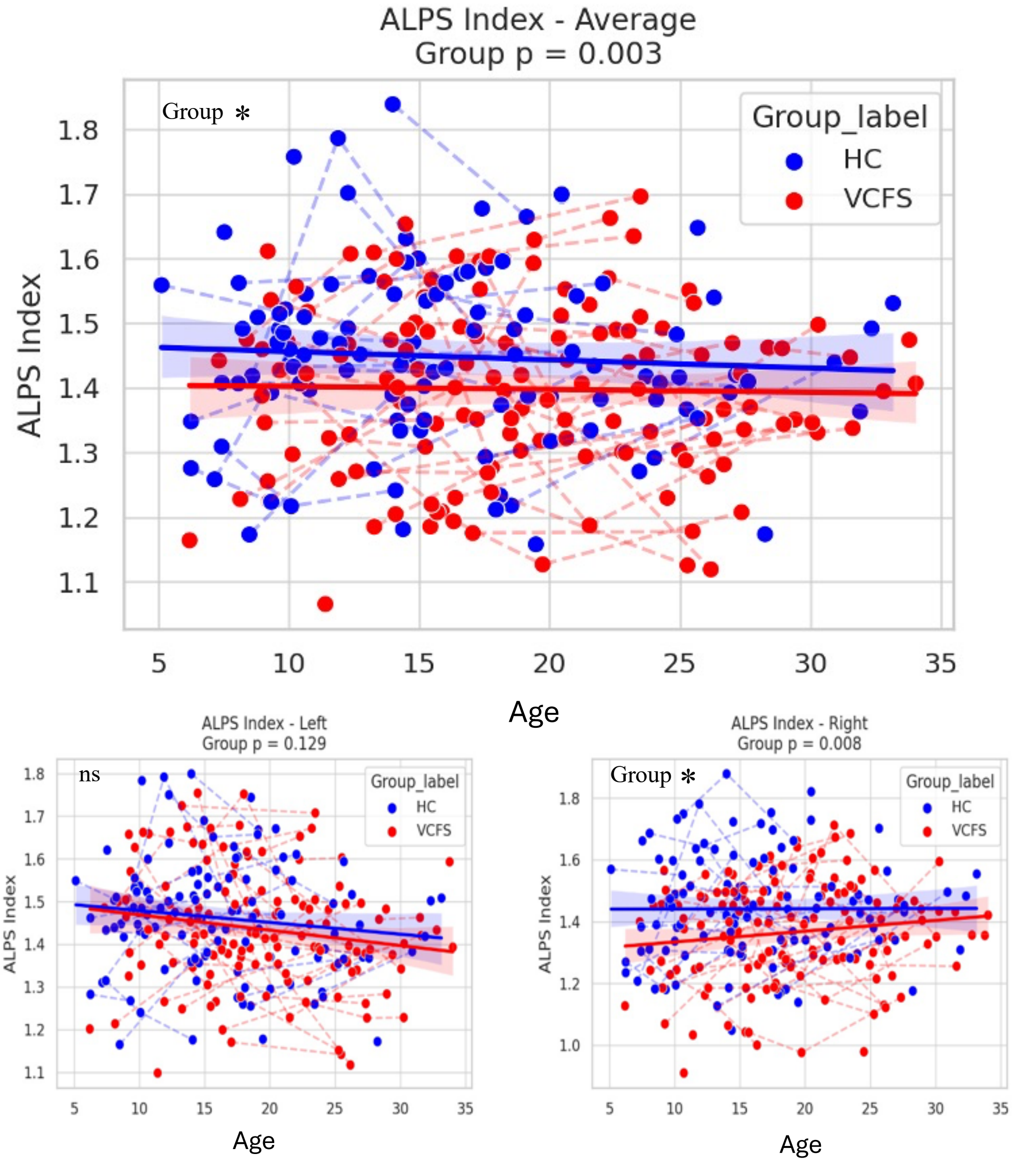
ALPS Index - Average
Group p = 0.003
Group *
Group_label
HC
VCFS
ALPS Index
Age
ALPS Index - Left
Group p = 0.129
ns
Group_label
HC
VCFS
ALPS Index
Age
ALPS Index - Right
Group p = 0.008
Group *
Group_label
HC
VCFS
ALPS Index
Age

### 4. SUPPLEMENTARY REFERENCES


1. Mancini V, Saleh MG, Delavari F, Bagautdinova J, Eliez S. Excitatory/Inhibitory Imbalance Underlies Hippocampal Atrophy in Individuals With 22q11.2 Deletion Syndrome With Psychotic Symptoms. Biol Psychiatry. 2023 Oct 1;94(7):569–79.

2. Saleh MG, Rimbault D, Mikkelsen M, Oeltzschner G, Wang AM, Jiang D, et al. Multi-vendor standardized sequence for edited magnetic resonance spectroscopy. NeuroImage. 2019 Apr 1;189:425–31.

3. Taoka T, Masutani Y, Kawai H, Nakane T, Matsuoka K, Yasuno F, et al. Evaluation of glymphatic system activity with the diffusion MR technique: diffusion tensor image analysis along the perivascular space (DTI-ALPS) in Alzheimer's disease cases. Jpn J Radiol. 2017 Apr;35(4):172–8.

4. Taoka T, Ito R, Nakamichi R, Nakane T, Sakai M, Ichikawa K, et al. Diffusion-weighted image analysis along the perivascular space (DWI–ALPS) for evaluating interstitial fluid status: age dependence in normal subjects. Jpn J Radiol. 2022 Sep 1;40(9):894–902.

5. Taoka T, Ito R, Nakamichi R, Nakane T, Kawai H, Naganawa S. Diffusion Tensor Image Analysis ALong the Perivascular Space (DTI-ALPS): Revisiting the Meaning and Significance of the Method. Magn Reson Med Sci. 2024 Apr 2;23(3):268–90.

6. Edden RAE, Puts NAJ, Harris AD, Barker PB, Evans CJ. Gannet: A batch-processing tool for the quantitative analysis of gamma-aminobutyric acid–edited MR spectroscopy spectra. J Magn Reson Imaging JMRI. 2014 Dec;40(6):1445–52.

7. Ashburner J, Friston KJ. Unified segmentation. NeuroImage. 2005 Jul 1;26(3):839–51.

8. Gasparovic C, Song T, Devier D, Bockholt HJ, Caprihan A, Mullins PG, et al. Use of tissue water as a concentration reference for proton spectroscopic imaging. Magn Reson Med. 2006 Jun;55(6):1219–26.

9. Wenneberg C, Nordentoft M, Rostrup E, Glenthøj LB, Bojesen KB, Fagerlund B, et al. Cerebral Glutamate and Gamma-Aminobutyric Acid Levels in Individuals at Ultra-high Risk for Psychosis and the Association With Clinical Symptoms and Cognition. Biol Psychiatry Cogn Neurosci Neuroimaging. 2020 Jun;5(6):569–79.

10. Saleh MG, Papantoni A, Mikkelsen M, Hui SCN, Oeltzschner G, Puts NA, et al. Effect of Age on GABA+ and Glutathione in a Pediatric Sample. AJNR Am J Neuroradiol. 2020 Jun;41(6):1099–104.

11. Conen S, Gregory CJ, Hinz R, Smallman R, Corsi-Zuelli F, Deakin B, et al. Neuroinflammation as measured by positron emission tomography in patients with recent onset and established schizophrenia: implications for immune pathogenesis. Mol Psychiatry. 2021 Sep;26(9):5398–406.
